# Isotretinoin use is associated with persistent alterations in male reproductive function

**DOI:** 10.64898/2026.07.22.26356991

**Authors:** Fie Bendix, Lærke Priskorn, Niels Jørgensen, Hanne Frederiksen, Anne Jørgensen, Lina Mørch, Hans Christian Ring, SriDurgaDevi Kolla, Anna K. Rosenmai, Terje Svingen, Anders Juul, Anders Rehfeld

**Author notes:** Corresponding author: (AR).

## Abstract

**Background:** Isotretinoin (ISO) is a systemic retinoid widely used for the treatment of *acne vulgaris*, yet its potential effects on male reproductive function remain unclear with previous studies reporting inconsistent findings.

**Methods and findings:** We conducted a multi-model study combining data from the Danish Young Men Study (DYMS) cohort with *ex vivo* cultured human testis tissue and an *in vitro* Retinoic Acid Receptor alpha (RARα) reporter gene assay. In the DYMS cohort semen parameters and reproductive hormone concentrations assessed on a specific examination day were compared between ISO-users (n=322) and non-users (n=4,065). *Ex vivo* human testis tissue was exposed to ISO and RA for 96 hours to assess changes in hormone secretion and germ cell dynamics, while the *in vitro* RARα assay evaluated ISO for effects on RA-induced RARα-signaling.

In the DYMS cohort, ISO use was associated with reduced FSH concentrations, higher inhibin B/FSH ratio, and elevated estradiol concentrations, along non-significant tendencies of lower sperm counts. These associations persisted even after cessation of ISO use but were not evident in men first initiating ISO use 1 year after their examination date. In the *ex vivo* testis tissue, ISO exposure altered hormone secretion, including increased testosterone, but did not affect germ cell proliferation or apoptosis. ISO also inhibited RA-induced RARα signaling *in vitro* in a competitive manner.

**Conclusions:** These findings suggests that ISO-use may influence male reproductive function, potentially with persistent effects. Further randomized clinical trials are needed to validate these effects.

## Introduction

Human sperm counts have declined globally by more than 50 % between 1973 and 2018 (1). Although the underlying causes remain unknown, emerging evidence indicate that exposure to reproductive toxicants may be implicated (2).

Isotretinoin (ISO) is a widely prescribed systemic drug for the treatment of moderate to severe *acne vulgaris* in young men and women (3). In 2024, 11,345 Danish men redeemed a prescription for ISO, with high usage observed among young men aged 18-24 years (14.54 per 1,000) (4,5). While ISO use is known to be associated with a range of adverse side effects (6), studies examining its effects on male reproductive function remain inconclusive with conflicting findings (7–22).

ISO, or 13-*cis*-retinoic acid, is a derivative of vitamin A and a stereoisomer of retinoic acid (RA) (23). *In vitro* studies have shown that ISO can activate the retinoid acid receptor alpha (RARα) at low nM concentrations, in a dose-dependent manner similar to that of RA (24). RARs are expressed in testis tissue, where RA-signaling has a vital role in the regulation of spermatogenesis (25,26). This regulatory role has been demonstrated both in animal models and humans, where disruption of RA-signaling leads to infertility due to impaired spermatogenesis (27–31). Accordingly, the RA-signaling pathway is currently exploited as a potential male contraceptive target (32,33), highlighting its essential role in male reproductive function.

Given the widespread use of ISO among young men, the importance of RA-signaling for spermatogenesis, and the current knowledge gap on the potential reproductive side effects of ISO, this study aimed to comprehensively investigate the effects of ISO on male reproductive function. To this end, we employed a multi-model approach compromising: 1) epidemiological association studies on ISO use, semen quality parameters, and reproductive hormone profiles in a cohort of young Danish men, 2) *ex vivo* studies in cultured human testis tissue exposed to ISO to assess hormone production and germ cell dynamics, and 3) an *in vitro* RARα transactivation model to test whether ISO interferes with RA-induced RARα-signaling.

## Materials and Methods

We employ a multi-model design that anchors on human epidemiological cohort data, with translational *ex vivo* experiments, and mechanistic *in vitro* assays providing complementary support within a harmonized analytical framework.

### Isotretinoin Use from Prescriptions in the DYMS cohort

#### Ethical Approval and Participants

All participants from the Danish Young Men Study (DYMS) cohort have given written informed consent, as approved by the regional scientific ethical committee of the Capital Region of Denmark with permit number H-K 289428. The men were compensated for their participation with 500 DKK (approximately 75 USD).

#### Study population

The DYMS cohort is a population of Danish young men (median age = 19) recruited when undergoing a compulsory physical examination for their suitability for military service, as previously described in detail (34,35). In brief, the men were approached on their military recruitment day by research staff from the Department of Growth and Reproduction, Rigshospitalet, Copenhagen, independently of their fit for the military. The men were invited to participate in a study aimed to understand reproductive function. When accepting study participation, the men were summoned for an examination day at Rigshospitalet where anthropometric and andrological physical examinations were performed. They also delivered a semen sample and had blood samples drawn.

The DYMS cohort was linked to the Danish National Patient Registry (DNPR) (36), the Danish CPR registry (37), and the Danish National Prescription Registry (NPR) (38,39) to assess prescription information of ISO for each participant. The NPR was established in 1994 and provides information on all individual prescriptions filled and redeemed at Danish pharmacies (38). Reporting to this registry is mandatory in Denmark and is performed automatically through standardized reporting systems in the pharmacies.

From 2002-2022, a total of 5,307 men were examined and accepted linking their data to the registries. For this study, we excluded men due to missing their exact examination day date (n=61) or abuse of anabolic steroids (n=29) thereby leaving a total of 5,217 men.

All ISO users were identified using the Anatomical Therapeutic Chemical Classification System (ATC) code linked to their specific redeemed prescriptions, categorized as D10BA01 “*systemic isotretinoin*”. ISO users were divided into three specific subgroups: 1) General ISO users (n=322), i.e., those that redeemed at least one ISO prescription at any timepoint prior to their examination; 2) Active ISO users (n=45), i.e., those with current ISO use on their examination day. Active ISO users were defined as having redeemed more than one ISO prescriptions in a 3-month window prior to their examination date, and minimum 14 days before the examination date; and 3) Prior ISO users (n=124), i.e., men who redeemed their last ISO prescription at least 2 years prior to their examination date. The three subgroups (and later ISO-users used for sensitivity analysis) are illustrated in figure 1. The subgroups are not mutually exclusive and overlap between the groups are found.

**Figure 1:**
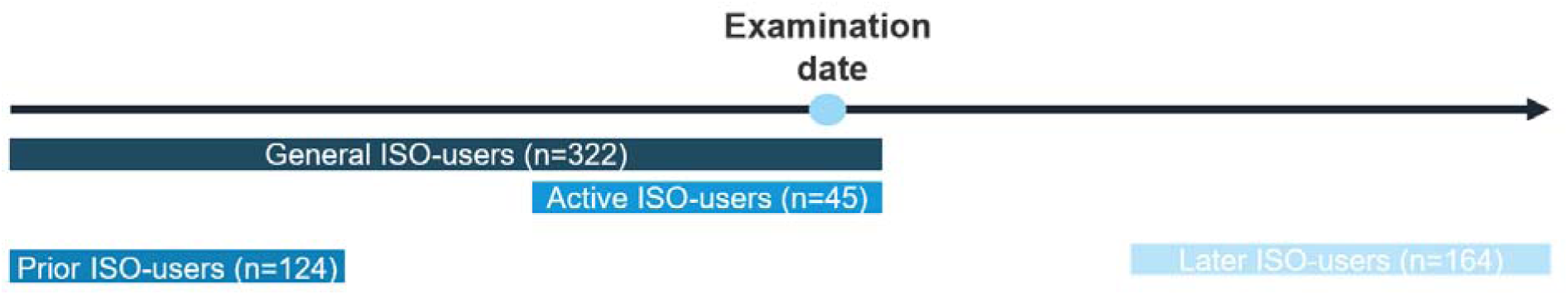
Graphical representation of ISO subgroups in the cohort. The subgroups of men using ISO in the DYMS cohort in relation to their individual examination day. The groups are not mutually exclusive and overlap between the groups in the analysis are displayed here. Number of participants in each sub-group are also shown. Abbreviations: ISO, isotretinoin; DYMS, Danish Young Men Study cohort.

The reference group was identified as non-ISO users with the exclusion of ISO at any point before their examination day (n=4,065). Also, a group of users of other acne medication (n=830) were identified by including the ATC-codes starting with D10 *“Anti-Acne Preparations”* (except D10BA01). This code includes both preparations for topical and systemic use, as a control for confounding by indication from the *acne vulgaris* disease itself.

Prescribed doses among ISO users were also retrieved from the NPR. The most common dose was 40 mg daily (n = 112), followed by 20 mg daily (n = 35), whereas few individuals were prescribed either 10 mg (n = 6) or 80 mg daily (n = 5). However, for most ISO users, the dose could not be deduced based on the information in the NPR (n = 164). Therefore, we analyzed all ISO users together as a single group, regardless of the available dosage information in the NPR.

#### Questionnaire

Before attending the examination day at the hospital, participants completed a standardized questionnaire. This collected information on current and past diseases, including genital conditions and fertility history, as well as lifestyle factors such as alcohol use, smoking habits, anabolic steroid use, physical activity, self-rated health, and maternal smoking during pregnancy. Information not available from registries was thus captured and could be adjusted for in the analyses.

#### Physical Examination

Weight and height of the men were measured, and body mass index (BMI) was calculated from the formula, weight (kg)/(height(m))^2^.

#### Semen Analysis

The semen samples were provided by masturbation during the morning hours of the examination day in a room adjacent to the hospital laboratory, following an advised abstinence period of at least 48 hours. The self-reported abstinence time was recorded. The samples were kept at 37°C until analysis. Analysis of semen was consistent throughout the study period and conducted in accordance with the current guidelines of the World Health Organization (WHO) (40), as previously described (34). Briefly total sperm count was calculated as semen volume (measured by weighing) multiplied by sperm concentration (million/ml, evaluated by hemocytometer). Sperm motility (%) was classified as immotile, locally motile, or progressively motile by manual microscopic examination. Total progressive motility count (TPMC) was calculated by total sperm count multiplied by the percentage of progressive motile sperm. Sperm morphology (%) was evaluated from a smear of the semen sample and stained with Papanicolaou.

#### Analysis of Serum Reproductive Hormone Concentrations

Blood samples were drawn from a cubital vein, and time of the blood sampling was recorded. The blood was then centrifuged, and the serum kept at −20°C until laboratory analysis of reproductive hormones which were performed in yearly batches at the Department of Growth and Reproduction, Copenhagen University Hospital – Rigshospitalet, Denmark. Serum concentrations of luteinizing hormone (LH), follicle-stimulating hormone (FSH), sex hormone-binding globulin (SHBG), and testosterone were assessed by time-resolved immunofluorometric assay (Delfia, Wallac, Turku, Finland). However, from 2014, testosterone and SHBG levels were determined using ELISA (Access2, Beckman Coultier Ltd, High Wycombe, UK), and from 2021 LH and FSH were assessed by immunoflurometric assay (Attelica, Siemens Healthineers, Erlangen, Germany). Estradiol was determined using radioimmunoassay (Pantex, Santa Monica, CA, USA). Finally, inhibin B levels were assessed using a two-sided enzyme immunometric assay (Serotec Microplate Reader, Kidlington, GB, or Tecan Sunrise Microplate Reader, Männedorf, Switzerland). Free testosterone was calculated from the measured total testosterone and SHBG with the equation and fixed albumin levels according to Vermeulen et al. (41). Ratios of testosterone/LH, and inhibin B/FSH were calculated as well.

#### Statistical Analysis

For the descriptive statistics the median (interquartile range (IQR), 25-75 pct) was calculated stratified by ISO use and use of other acne medication for the basic characteristics and for the markers of reproductive and testicular function.

For this cross-sectional analysis, adjusted linear regression analyses were conducted on the semen parameters and hormone concentrations to assess differences between ISO users at different time points, users of other acne medication, and, as the reference group, ISO non-users. The semen parameters and reproductive hormones were included as continuous variables and were transformed accordingly to pass the model assumption of linearity, normally distributed residuals, and homoscedasticity. Thus. semen volume, sperm concentration, and total sperm count were cubic-root-transformed, progressive sperm motility was squared, and morphology was square root transformed. All hormones were transformed to their natural logarithm.

Analyses were adjusted for potential confounders as well as variables that are associated with the precision of the outcome measurements. All analyses were adjusted for BMI (as categorical data divided into four categories: <18.5, 18.5-24.99, 25-30, <30), and age (as continuous data). Regression analyses for semen parameters were further adjusted for abstinence time and time from sample delivery to analyses (as continuous data), while reproductive hormones concentrations were adjusted for time of blood sampling (as continuous data). Adjusted β-coefficients with corresponding 95% confidence intervals and p-values were estimated. *P*-values < 0.05 were considered statistically significant.

All statistical analyses were conducted in R (R version 4.4.3, Vienna, Austria).

#### Sensitivity analysis

The adjusted linear regression analyses were repeated in a subgroup of later ISO users, i.e., men initiating ISO use at least one year after their examination date (figure 1) to ensure that the observed effects were solely from ISO use and not present but undiagnosed or untreated acne. The one-year margin was applied to eliminate the possibility of ISO use before or during the examination day.

#### Data availability

The datasets are not publicly available due to national data security legislation but are available from the corresponding author on reasonable request and with permission from the ethical committee of the capital region of Denmark.

### Isotretinoin and Retinoic Acid Exposure in *Ex Vivo* Cultured Testis Tissue

#### Ethical Approval, Participants and Human Testis Tissue Collection

The testis tissue used for culture was collected from testicular cancer patients recruited from the Andrology Clinic of the Department of Growth and Reproduction, Rigshospitalet, providing informed consent before their orchidectomy, as approved by the regional scientific ethical committee of the Capital Region of Denmark with permit number H-1-2012-007 in compliance with the Helsinki Declaration. In the period from September 2023 to December 2024, testis tissue from 11 patients undergoing orchidectomy was collected. For this study, experiments from 8 donors were included. The reasons for the exclusion of the remaining 3 donors were changes in the experimental setup in the beginning of the study, the amount of tissue received after surgical removal, and low tissue quality. After surgical removal at Copenhagen University Hospital - Rigshospitalet, the testis was immediately transported to the in-house Pathology Department, and macroscopically divided into tumor and non-tumor tissue pieces. A minimal portion of the non-tumor tissue was assigned for research and biobank use, while the majority of tissue was used for diagnostic purposes. The non-tumor testis tissue was used for laboratory experiments on the same day of the orchidectomy and set in culture within 3 hours of the surgical removal. Importantly, the non-tumorous tissue used for culture all originated from patients with testicular cancer. The tumor tissue, which was not cultured, was later histologically evaluated by a clinician: of the 8 donors included, 7 were diagnosed with germ cell tumors of the seminoma type (Donor 1, 3, 4, 5, 6, 7, and 8) and one with a spermatocytic tumor (Donor 2). Tissue from donor 1 was not exposed to the full panel of ISO and RA concentrations.

#### Sample Preparation and treatment concentrations

Testis tissue assigned for research was rapidly transported to the laboratory in culture medium containing DMEM/F-12 medium supplemented with 1% Penicillin-Streptomycin (Pen-Strep) (10,000 U/mL), 10% Fetal Bovine Serum (FBS), and 1% Insulin-Transferrin-Selenium-Sodium Pyruvate (ITS-A) (all Gibco™, NY, USA), and prepared for *ex vivo* culture as previously described (42). Briefly, within 3 hours of surgical removal, the tissue was dissected into approximately 1 mm³ fragments and cultured individually in 40 µL hanging droplets containing culture medium with treatment compounds or as vehicles. For each treatment group, 9 tissue fragments were cultured per Petri dish, with two technical replicates (18 fragments per treatment in total). The treatment compounds, ISO (Supelco, PA, USA) and RA (Sigma-Aldrich, MO, USA), were stored dry at −20°C and dissolved in dimethyl sulfoxide (DMSO) (Sigma-Aldrich, MO, USA) on the day of culture setup to prepare 10 mM stock solutions, which were protected from light until use.

To investigate the effects of ISO and compare these with the effects of RA, explants were cultured for a relative short period of 96 hours at 34°C and 5% CO in the presence of vehicle controls (0.1% or 0.25% DMSO), 1 µM RA, 10 µM RA, 10 µM ISO, 15 µM ISO, 25 µM ISO, or a combination of 10 µM RA+10 µM ISO. Media were changed after 48 hours, and culture media from the two technical replicates were pooled and collected after 48 and 96 hours for later hormone analyses. After the completed culture period of 4 days (96 hours), the 9 tissue fragments in each replicate from each treatment group were collected and distributed for analysis: Survival Assay (2 fragments), FFPE and immunohistochemistry (IHC), including the Proliferation Assay (4 fragments), or stored at −80℃ for potential additional genomic or proteomic analyses (3 fragments).

#### Survival Assay

To assess the presence of metabolically active cells in tissue fragments following *ex vivo* culture, a survival assay was performed using the resazurin-based In Vitro Toxicology Assay Kit (TOX-8, Sigma-Aldrich, Darmstadt, Germany) as previously described (42). Viable cells reduce resazurin (blue) to resorufin (pink), indicating aerobic metabolic activity. Color change was evaluated qualitatively and macroscopically. Only the tissue fragments within each treatment and replicate showing aerobic metabolic activity were further analyzed (Data not shown but available on request).

#### Immunohistochemistry

FFPE and IHC was performed on *ex vivo* cultured testis tissue, as previously described (42). Briefly, tissue pieces from each treatment condition were fixed in formalin (30 min at room temperature and overnight at 4 °C), embedded in paraffin, and sectioned using a rotational microtome (HM355S automatic, Thermo Scientific, MA, USA) onto Superfrost glass slides (Thermo Scientific, MA, USA).

Sections were deparaffinized, rehydrated, and subjected to antigen retrieval by microwaving in retrieval buffer (specified in Table 1). Endogenous peroxidase activity was blocked using H□O□, followed by blocking in 0.5% milk powder in TBS. Sections were incubated with primary antibodies (overnight at 4 °C) and corresponding secondary antibodies using the ImmPRESS polymer kit (Vector Laboratories, CA, USA) (Specified in Table 1). Signal was visualized with the ImmPACT AEC substrate kit (Vector Laboratories, CA, USA) and counterstained with Mayer’s hematoxylin. Slides were mounted with Aquamount (Merck, Darmstadt, Germany).

**Table 1:** Antibodies and conditions used for IHC.

| Antibody | Species | Dilution | Retrieval buffer | Antibody supplier and product number |
| --- | --- | --- | --- | --- |
| D2-40 | Mouse | 1:250 | Citrate | Dako M3619 |
| c-PARP | Rabbit | 1:400 | Citrate | Cell signaling 5625 |
| BrdU | Mouse | 1:400 | TEG | Invitrogen 15297397 |

Abbreviation: c-PARP, cleaved poly (ADP-ribose) polymerase; IHC, immunohistochemistry; TEG, Citrate buffer: 10nM, pH 6.0; TEG byffer: 10nM Tris, 0.5mM EGTA, pH 9.0

Appropriate positive and negative controls were included, and only sections with expected control staining were analyzed. Slides were scanned using a NanoZoomer 2.0 HT (Hamamatsu Photonics, Shizuoka, Japan) and evaluated qualitatively in NDP.view2 software (Hamamatsu Photonics). Semiquantitative assessment was performed using a predefined scoring system, as described in (42), and the QuPath v0.5.1 software (43) to estimate positive cell ratio per tubule. As no germ cell-specific antibody was included in the IHC panel, cells were classified based on their localization within seminiferous tubules and morphology rather than confirmed IHC germ cell identity.

As the tissue originated from patients with testicular cancer, immunostaining for D2-40, a marker of germ cell neoplasia in situ (GCNIS), was performed. GCNIS-positive seminiferous tubules were detected in tissue from all donors except donor 2, who was diagnosed with a spermatocytic tumor. Overall, 35 of 88 tissue fragments (39.8%) contained one or more GCNIS-positive tubules (representative histological images from each donor are provided in supplementary figure S1).

#### Proliferation Assay

Proliferation of germ cells was assessed using a 5-Bromo-2-deoxyuridine (BrdU) incorporation assay, as previously described (42). BrdU labelling reagent (Invitrogen™, CA, USA) was added to the culture medium (1:10 dilution) and incubated with testis tissue fragments during the last 6 hours of the 96 hours culture period. Tissue fragments were subsequently fixed and embedded in paraffin as described above. BrdU incorporation was visualized by IHC using a BrdU-specific antibody (Specified in table 1), and cells showing positive nuclear staining were considered proliferating.

#### Steroid Hormone Analyses by Liquid Chromatography-tandem Mass Spectrometry (LC-MS/MS)

Steroid hormones were measured in the stored culture media using two different sensitive isotope-diluted TurboFlow-LC-MS/MS methods clinically validated for eighteen steroid metabolites in human serum (44,45) at the Department of Growth and Reproduction, Copenhagen University Hospital – Rigshospitalet, Denmark, and further modified for measurement in the culture media. These analytical methods included a comprehensive range of steroid hormones; the estrogens (estrone and estradiol) and androgens (dehydroepiandrosterone, dehydroepiandrosterone sulphate (DHEAS), androstenedione (adion), testosterone and dihydrotestosterone (DHT)) as well as the key steroidogenic intermediates (17-hydroxyprognenolone, 17-hydroxyprogesterone (17-OHP), progesterone), mineralocorticoids (11-deoxycorticosterone (DOC), corticosterone, aldosterone) and glucocorticoids (11-deoxycortisol (11-DOC), cortisol and cortisone). The samples from the culture media were measured for most of the steroid metabolites in 2 batches in spring 2024 and winter 2025 including approximately 40 experimental samples in each batch. For the estrogens, a third batch containing the same samples as the two previous batches were measured in winter 2026. For all batches, calibration standards prepared in culture media, one blank (water), and a set of method controls comprising unspiked media, media spiked with low and high concentrations of native steroid standards and well-known pooled control samples were used. Calibration curves were generated using media-based standards to account for matrix effects. Prior to analysis, all collected media samples were diluted fourfold in culture media followed by addition of a mixture of isotope labeled steroid standards to all samples, calibration and control material. In a subset of samples, an additional dilution was performed to enable reanalysis of testosterone concentrations within optimal range. This approach ensured consistency and reliability across batches and concentration ranges.

For all analytical batches included in this study, the relative standard deviation (RSD) was <5.5% for steroid metabolites in low spike levels, whereas RSD was <3.6% for steroid metabolites in the controls spiked at high level.

#### Inhibin B Detection by ELISA

Inhibin B was detected in the stored culture media by ELISA (Beckman Coulter Inhibin B Gen II ELISA, Beckman Coulter, Brea, CA, USA). The samples were diluted 1:10 with culture media, with some samples requiring a 1:20 dilution, and measured in three batches in winter 2024, spring 2024, and winter 2025 including approximately 30-40 experimental samples in each batch. The detection limit of the Inhibin B assay was 3 ng/L and inter-assay variation was <11%.

#### Statistical Analysis

The quantified positive cell ratio from the proliferation assay and the measured concentrations from the hormone analysis were evaluated using multiple linear regression models in R (version 4.4.1) and RStudio (version 2024.09.1). To pass the model assumptions of linearity, normally distributed residuals, and homoscedasticity the data from the treatment media measurements was all logarithmic transformed. Donor and time (48 hours vs. 96 hours of culture in the saved treatment media) effects were included to account for inter-individual variability. P-values <0.05 were considered statistically significant, and denoted as followed; *p<0.05, **p<0.01, ***p<0.001. Plots were generated using RStudio.

#### Data availability

Data can be shared upon request.

### Isotretinoin and Retinoid Acid’s Effects on RARα Reporter Gene Assay

The human RARα reporter (Luc)-HEK293 cell line (BPS Bioscience, CA, USA) was used. Cells were cultured in high glucose DMEM with phenol red (Cytiva, Bronshoj, Denmark) supplemented with 10 % FBS (Gibco, MA, USA), 50 units/mL penicillin (Gibco, MA, USA), 50 μg/mL streptomycin (Gibco, MA, USA), 400 μg/L geneticin (Gibco, MA, USA), 1 μg/mL puromycin (Sigma Aldrich, MO, USA) and 100 μg/mL hygromycin (Invitrogen, CA, USA). Assay medium was DMEM without phenol red (Gibco, MA, USA) supplemented with 10 % DCC-FBS (Cytivia, Bronshoj, Denmark), 50 units/mL penicillin, and 50 μg/mL streptomycin (Gibco, MA, USA). Cells were maintained at 37 °C and 5 % CO_2_ in a humidified atmosphere.

Cells were seeded at 40,000 cells/well in white 96-well plates (Revity, MA, USA) and left for 22-24 h at 37 °C and 5 % CO_2_ in a humidified atmosphere. Successively, cells were exposed to RA (Sigma, MO, USA) and ISO (Sigma, MO, USA) for approximately 24 h. RA was tested at concentrations ranging from 0.015-100 nM with background of 0, 12, 195, or 1440 nM ISO for approximately 24 h. ISO concentrations were chosen based on its EC50 value for RARα activation (12nM) (24) and its plasma C_max_ in humans (1440 nM) (46). Background concentrations of DMSO were kept constant across plates at 0.1%. After exposure, luminescence was measured using Bright-Glo™ Luciferase Assay System (Promega, WI, USA) following the protocol provided by the manufacturer.

Experiments were repeated on three different days and included 3 technical replicates per treatment within each repeat. Data are presented as means of absolute luminescence where data from each independent experiment was pooled

#### Data availability

Data can be shared upon request.

## Results

### Basic characteristics of the young men

The basic characteristics of the included young men are presented in Table 2, stratified by reference group, and users of acne medication: either ISO (the “general ISO users”) or other acne medication. The median age was 19 years for all three groups, and BMI of 22.2, 22.5, and 22.4 kg/m^2^, respectively. The median abstinence time was 61, 61, and 62 hours, with a delay of time from ejaculation to analyses of motility 33, 35, and 33 minutes, respectively. Blood samples for all three groups were collected at a standardized timeframe in the morning of the examination day with medians of 09:55, 09:55, and 10:00, respectively. In the reference group, 49% reported occasional or daily cigarette smoking. Among medication users, the prevalence was slightly lower, with 42% for ISO users and 46% for users of other acne medications. Still, these proportions remain broadly comparable to those of the reference group. Consequently, no differences were observed between the groups, indicating that they are comparable for subsequent analyses.

**Table 2:** Basic characteristics of the young men stratified by group.

|  | Reference group | Acne medication users |  |
| --- | --- | --- | --- |
|  | Non-users<br>N = 4,065<br>Median [25-75 pct] | Isotretinoin (ISO)<br>N = 322<br>Median [25-75 pct] | Other acne medication<br>N = 830<br>Median [25-75 pct] |
| **Basic characteristics** |  |  |  |
| Age, years | 19.0 [18.7, 19.6] | 19.0 [18.7, 19.5] | 19.0 [18.7, 19.6] |
| BMI, kg/m<sup>2</sup> | 22.2 [20.5, 24.2] | 22.5 [20.5, 24.4] | 22.4 [20.7, 24.2] |
| Height, cm | 182 [177, 186] | 182 [178, 187] | 182 [178, 187] |
| Weight, kg | 74 [67, 81] | 74 [68, 81] | 74 [68, 81] |
| Abstinence time, hours | 61 [58, 83] | 61 [57, 82] | 62 [57, 84] |
| Delay, minutes | 33 [25, 49] | 35 [23, 50] | 33 [23, 50] |
| Time of blood sampling, hh:mm | 09:55 [09:25, 10:30] | 09:55 [09:20, 10:30] | 10:00 [09:25, 10:35] |
| Smoking status, yes (%) | 1,966 (49%) | 134 (42%) | 377 (46%) |

Data are illustrated as median (25-75 percentile). Other “acne medication” is defined from ATC-codes “D10 Anti-acne preparations” except systemic “D10BA01 Isotretinoin” as described in the Methods section. Abbreviation: BMI, body mass index.

### Semen parameters and hormone concentrations in young men using isotretinoin

The median sperm concentration, total sperm count, and total number of progressively motile spermatozoa were 41 mill/ml, 125 mill and 74 mill among general ISO users and 45 mill/ml, 145 mill, and 88 mill among non-users, respectively (Table 3), however the differences did not reach statistical significance. In contrast, this pattern was not evident among users of other acne medication, whose median sperm concentration, total sperm count, and total number of progressively motile spermatozoa were 45 mill/ml, 141 mill and 90 mill, respectively, and thereby largely comparable to those of the non-users (Table 3). For the ISO-users that actively used ISO during their examination day, the corresponding figures were 41 mill/ml, 115 mill and 68 mill, and for the ISO users that had ceased treatment at least two years prior to participation in the examination day the corresponding figures were 39 mill/ml, 119 mill and 74 mill, however, also non-significant compared to non-users (Table 4).

**Table 3:**
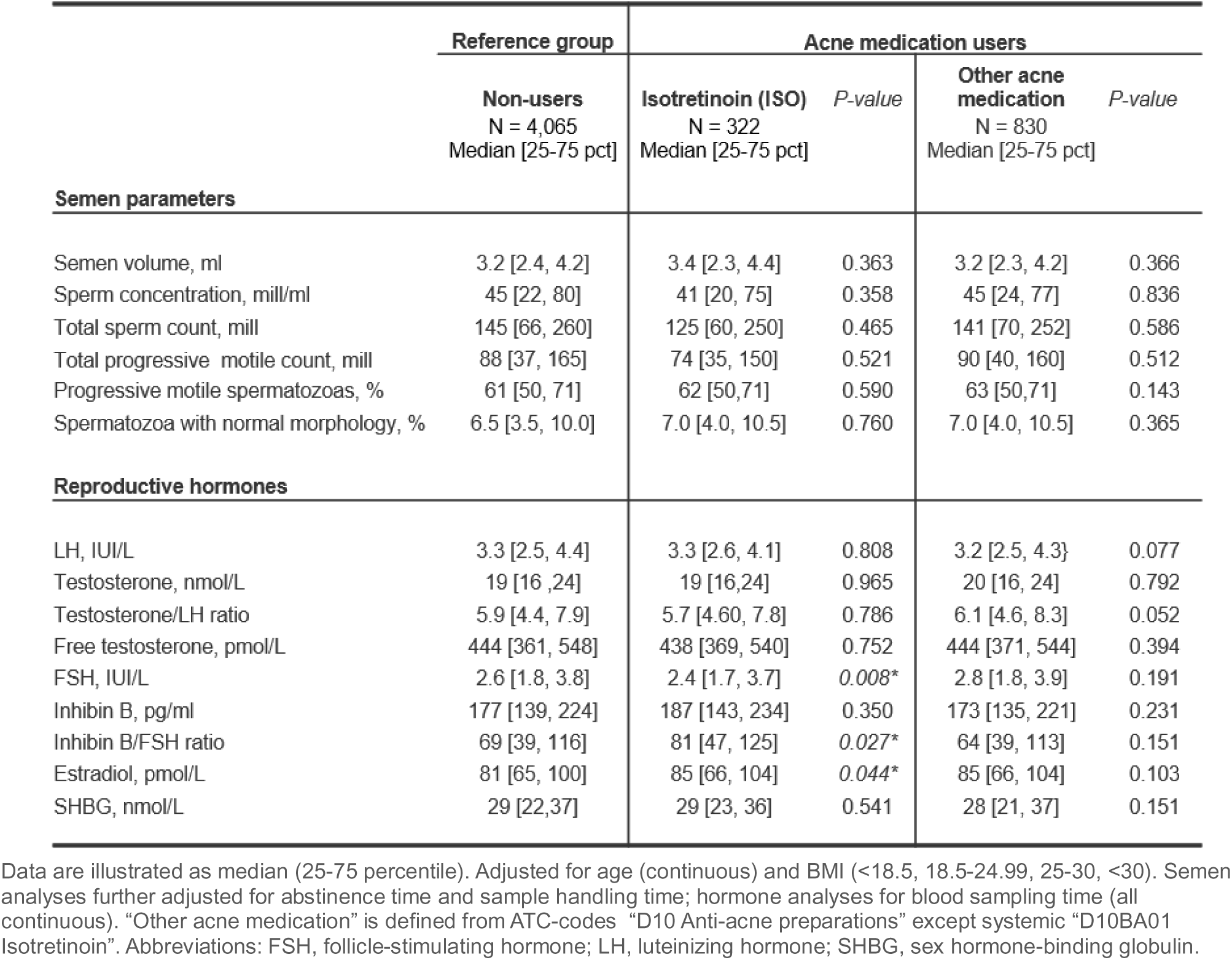
Use of acne medication in young men at any timepoint prior the examination date.

|  | Reference group | Acne medication users |  |  |  |
| --- | --- | --- | --- | --- | --- |
|  | Non-users<br>N = 4,065<br>Median [25-75 pct] | Isotretinoin (ISO)<br>N = 322<br>Median [25-75 pct] | P-value | Other acne medication<br>N = 830<br>Median [25-75 pct] | P-value |
| **Semen parameters** |  |  |  |  |  |
| Semen volume, ml | 3.2 [2.4, 4.2] | 3.4 [2.3, 4.4] | 0.363 | 3.2 [2.3, 4.2] | 0.366 |
| Sperm concentration, mill/ml | 45 [22, 80] | 41 [20, 75] | 0.358 | 45 [24, 77] | 0.836 |
| Total sperm count, mill | 145 [66, 260] | 125 [60, 250] | 0.465 | 141 [70, 252] | 0.586 |
| Total progressive motile count, mill | 88 [37, 165] | 74 [35, 150] | 0.521 | 90 [40, 160] | 0.512 |
| Progressive motile spermatozoas, % | 61 [50, 71] | 62 [50, 71] | 0.590 | 63 [50, 71] | 0.143 |
| Spermatozoa with normal morphology, % | 6.5 [3.5, 10.0] | 7.0 [4.0, 10.5] | 0.760 | 7.0 [4.0, 10.5] | 0.365 |
| **Reproductive hormones** |  |  |  |  |  |
| LH, IU/L | 3.3 [2.5, 4.4] | 3.3 [2.6, 4.1] | 0.808 | 3.2 [2.5, 4.3] | 0.077 |
| Testosterone, nmol/L | 19 [16, 24] | 19 [16, 24] | 0.965 | 20 [16, 24] | 0.792 |
| Testosterone/LH ratio | 5.9 [4.4, 7.9] | 5.7 [4.60, 7.8] | 0.786 | 6.1 [4.6, 8.3] | 0.052 |
| Free testosterone, pmol/L | 444 [361, 548] | 438 [369, 540] | 0.752 | 444 [371, 544] | 0.394 |
| FSH, IU/L | 2.6 [1.8, 3.8] | 2.4 [1.7, 3.7] | 0.008* | 2.8 [1.8, 3.9] | 0.191 |
| Inhibin B, pg/ml | 177 [139, 224] | 187 [143, 234] | 0.350 | 173 [135, 221] | 0.231 |
| Inhibin B/FSH ratio | 69 [39, 116] | 81 [47, 125] | 0.027* | 64 [39, 113] | 0.151 |
| Estradiol, pmol/L | 81 [65, 100] | 85 [66, 104] | 0.044* | 85 [66, 104] | 0.103 |
| SHBG, nmol/L | 29 [22, 37] | 29 [23, 36] | 0.541 | 28 [21, 37] | 0.151 |

Data are illustrated as median (25-75 percentile). Adjusted for age (continuous) and BMI (<18.5, 18.5-24.99, 25-30, >30). Semen analyses further adjusted for abstinence time and sample handling time; hormone analyses for blood sampling time (all continuous). "Other acne medication" is defined from ATC-codes "D10 Anti-acne preparations" except systemic "D10BA01 Isotretinoin". Abbreviations: FSH, follicle-stimulating hormone; LH, luteinizing hormone; SHBG, sex hormone-binding globulin.

**Table 4:**
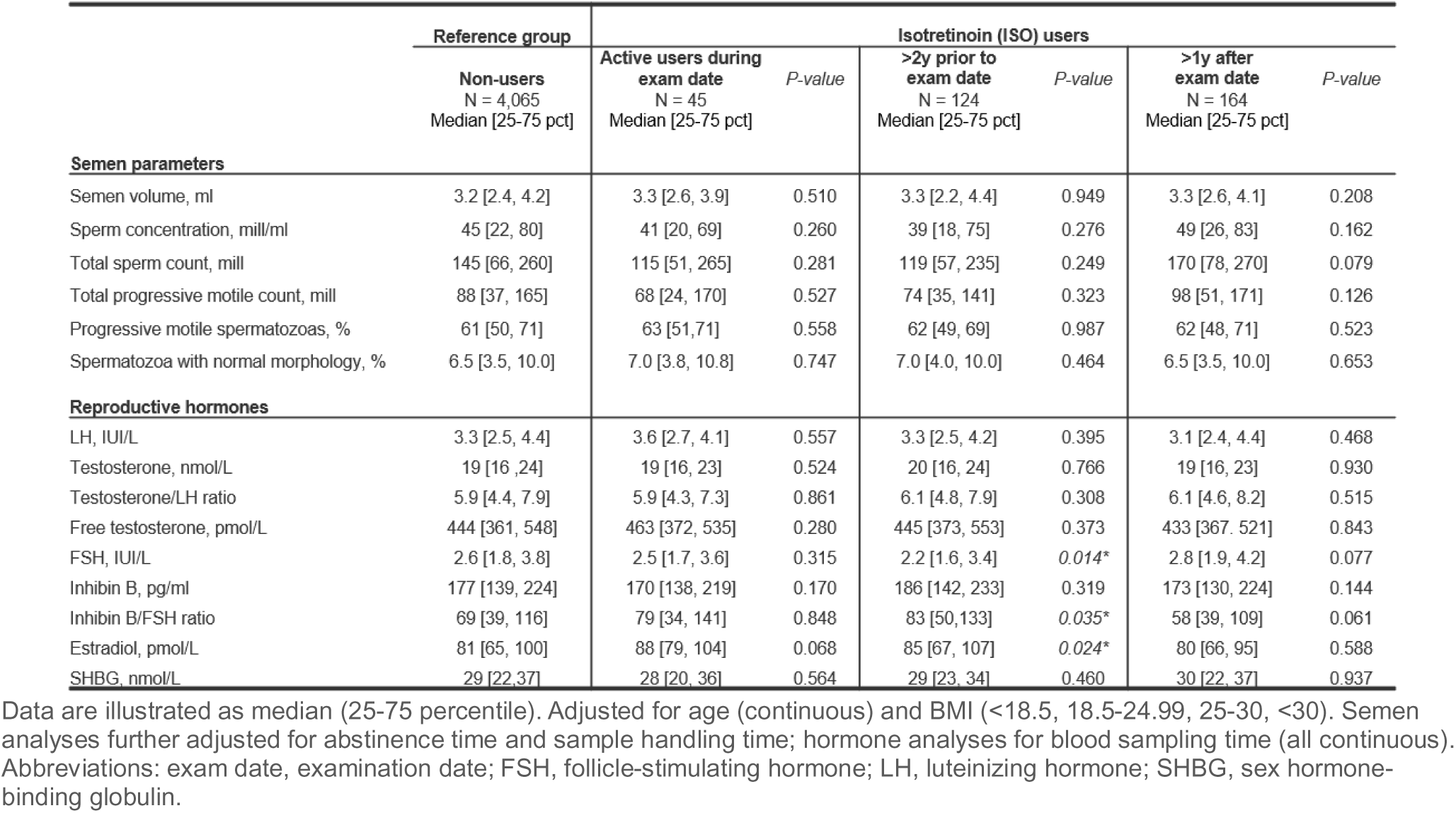
ISO use in young men during, prior, or after their examination date.

|  | Reference group | Isotretinoin (ISO) users |  |  |  |  |  |
| --- | --- | --- | --- | --- | --- | --- | --- |
|  | Non-users<br>N = 4,065<br>Median [25-75 pct] | Active users during<br>exam date<br>N = 45<br>Median [25-75 pct] | P-value | >2y prior to<br>exam date<br>N = 124<br>Median [25-75 pct] | P-value | >1y after<br>exam date<br>N = 164<br>Median [25-75 pct] | P-value |
| **Semen parameters** |  |  |  |  |  |  |  |
| Semen volume, ml | 3.2 [2.4, 4.2] | 3.3 [2.6, 3.9] | 0.510 | 3.3 [2.2, 4.4] | 0.949 | 3.3 [2.6, 4.1] | 0.208 |
| Sperm concentration, mill/ml | 45 [22, 80] | 41 [20, 69] | 0.260 | 39 [18, 75] | 0.276 | 49 [26, 83] | 0.162 |
| Total sperm count, mill | 145 [66, 260] | 115 [51, 265] | 0.281 | 119 [57, 235] | 0.249 | 170 [78, 270] | 0.079 |
| Total progressive motile count, mill | 88 [37, 165] | 68 [24, 170] | 0.527 | 74 [35, 141] | 0.323 | 98 [51, 171] | 0.126 |
| Progressive motile spermatozoas, % | 61 [50, 71] | 63 [51, 71] | 0.558 | 62 [49, 69] | 0.987 | 62 [48, 71] | 0.523 |
| Spermatozoa with normal morphology, % | 6.5 [3.5, 10.0] | 7.0 [3.8, 10.8] | 0.747 | 7.0 [4.0, 10.0] | 0.464 | 6.5 [3.5, 10.0] | 0.653 |
| **Reproductive hormones** |  |  |  |  |  |  |  |
| LH, IU/L | 3.3 [2.5, 4.4] | 3.6 [2.7, 4.1] | 0.557 | 3.3 [2.5, 4.2] | 0.395 | 3.1 [2.4, 4.4] | 0.468 |
| Testosterone, nmol/L | 19 [16, 24] | 19 [16, 23] | 0.524 | 20 [16, 24] | 0.766 | 19 [16, 23] | 0.930 |
| Testosterone/LH ratio | 5.9 [4.4, 7.9] | 5.9 [4.3, 7.3] | 0.861 | 6.1 [4.8, 7.9] | 0.308 | 6.1 [4.6, 8.2] | 0.515 |
| Free testosterone, pmol/L | 444 [361, 548] | 463 [372, 535] | 0.280 | 445 [373, 553] | 0.373 | 433 [367, 521] | 0.843 |
| FSH, IU/L | 2.6 [1.8, 3.8] | 2.5 [1.7, 3.6] | 0.315 | 2.2 [1.6, 3.4] | 0.014* | 2.8 [1.9, 4.2] | 0.077 |
| Inhibin B, pg/ml | 177 [139, 224] | 170 [138, 219] | 0.170 | 186 [142, 233] | 0.319 | 173 [130, 224] | 0.144 |
| Inhibin B/FSH ratio | 69 [39, 116] | 79 [34, 141] | 0.848 | 83 [50, 133] | 0.035* | 58 [39, 109] | 0.061 |
| Estradiol, pmol/L | 81 [65, 100] | 88 [79, 104] | 0.068 | 85 [67, 107] | 0.024* | 80 [66, 95] | 0.588 |
| SHBG, nmol/L | 29 [22, 37] | 28 [20, 36] | 0.564 | 29 [23, 34] | 0.460 | 30 [22, 37] | 0.937 |

Data are illustrated as median (25-75 percentile). Adjusted for age (continuous) and BMI (<18.5, 18.5-24.99, 25-30, >30). Semen analyses further adjusted for abstinence time and sample handling time; hormone analyses for blood sampling time (all continuous). Abbreviations: exam date, examination date; FSH, follicle-stimulating hormone; LH, luteinizing hormone; SHBG, sex hormone-binding globulin.

For the reproductive hormones, FSH was lower and inhibin B/FSH ratio higher for general ISO users vs. non-users, 2.4 vs. 2.6 IUI/L, and 81 vs. 69, respectively (Table 3). Notably, the inhibin B/FSH ratio increase does not appear to be driven by decreased FSH alone, but also by an insignificant increased in inhibin B. For LH, testosterone, and SHBG no tendencies were observed, while estradiol was significantly higher among ISO users compared to non-users, 85 vs. 81 pmol/L (Table 3). No significant associations were seen for users of other acne medication (Table 3). The active ISO users also tended to have higher estradiol concentrations, but not significant (P=0.06, Table 4). However, the prior ISO users, as the general ISO users, had lower FSH, higher inhibin B/FSH ratio, and higher estradiol, when compared to non-users, 2.2 vs. 2.6 IUI/L, 83 vs. 69, and 85 vs. 81 pmol/L, respectively (Table 4).

As a sensitivity analysis we examined the subgroup of young men who first initiated ISO use at least 1 year after their examination date (Table 4). In this subgroup, no significant differences were observed, nor the tendencies seen for hormone or semen parameters among the other groups of ISO-users.

### Hormone concentrations in cultured human testis tissue exposed to isotretinoin

Given that our epidemiological analysis revealed associations between ISO use and reproductive hormone levels, we next sought to determine whether these hormonal changes were reflected in cultured human testis tissue exposed to ISO.

To evaluate if ISO and RA affected the hormone secretion in the cultured testis samples, concentrations of secreted androgens, estrogen, and inhibin B were quantified from the treatment media using LC-MS/MS and ELISA. Testosterone concentrations were elevated across all treatment conditions compared to the vehicle control, except for 1µM RA (Figure 2a). In contrast, DHT concentrations were reduced after all treatment conditions except 1µM RA (Figure 2b). In the testis, testosterone can undergo conversion to estradiol by aromatase but none of the ISO or RA treatments altered estradiol concentrations (Figure 2c). Inhibin B concentrations were lower for 25 µM ISO and 10 µM RA+10 µM ISO (Figure 2d). Additional intermediate metabolites involved in steroid biosynthesis were also measured. These data, along with raw hormone concentrations, are included in the supplementary materials (Supplementary figures, S2-4).

**Figure 2:**
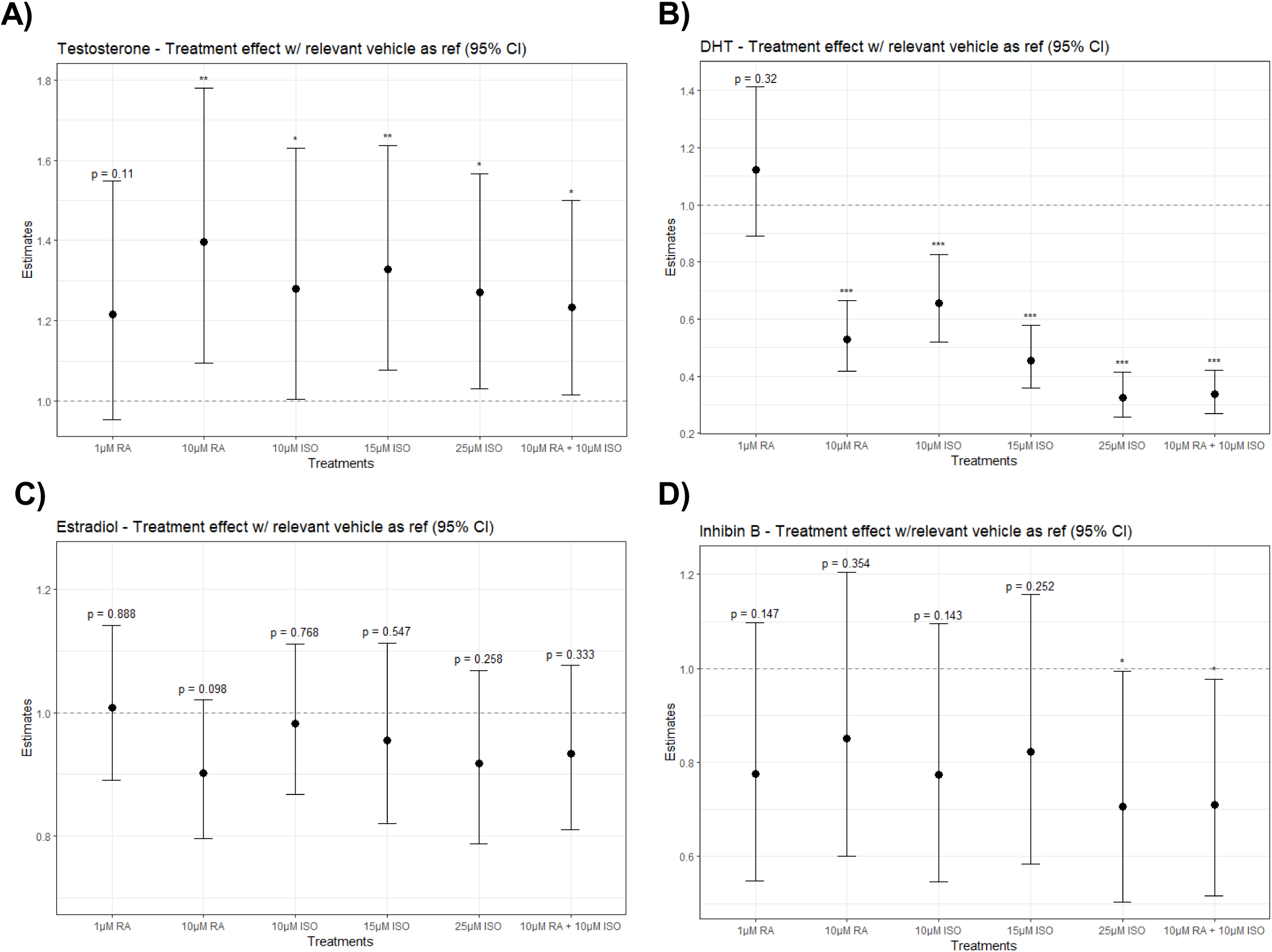
Effect on selected steroid hormones and inhibin B after ISO or RA exposure in cultured human testis tissue. The effect of different RA and ISO concentrations (x-axis) on cultured human testis tissue on A) testosterone, B) DHT, C) estradiol, and D) inhibin B levels (n=8). The estimates (y-axis) are created with multiple linear regression on log-transformed hormone concentrations to their corresponding vehicle control (either 0.1% or 0.25% DMSO). CI of 95%. Dashed line represents the line of null effect. *p<0.05, **p<0.01, ** 0.001. Abbreviations: CI, Confidence Intervals; RA, Retinoic Acid; ISO, Isotretinoin; DHT, Dihydrotestosterone.

### Changes in the germ cell dynamics in ex vivo cultured testis tissue after isotretinoin exposure

As a tendency towards reduced sperm concentration and sperm count was observed in the young men after ISO use, we next investigated whether this could be reflected by alterations in germ cell proliferation and apoptosis within the *ex vivo* cultured human testis tissue. As no germ cell-specific marker was included, the identity of germ cells was based on their localization and morphology within the seminiferous tubules.

Proliferation (premeiotic mitotic division) was assessed by BrdU incorporation in a subset of presumed germ cells (spermatogonia) and quantified in each seminiferous tubule (Figure 3a, upper panel). The fraction of BrdU-positive cells relative to all putative germ cells in each tubule was quantitatively assessed, and the effect of each treatment is presented relative to its vehicle control (Figure 3b). No effect of ISO or RA treatment was observed on cell proliferation.

**Figure 3:**
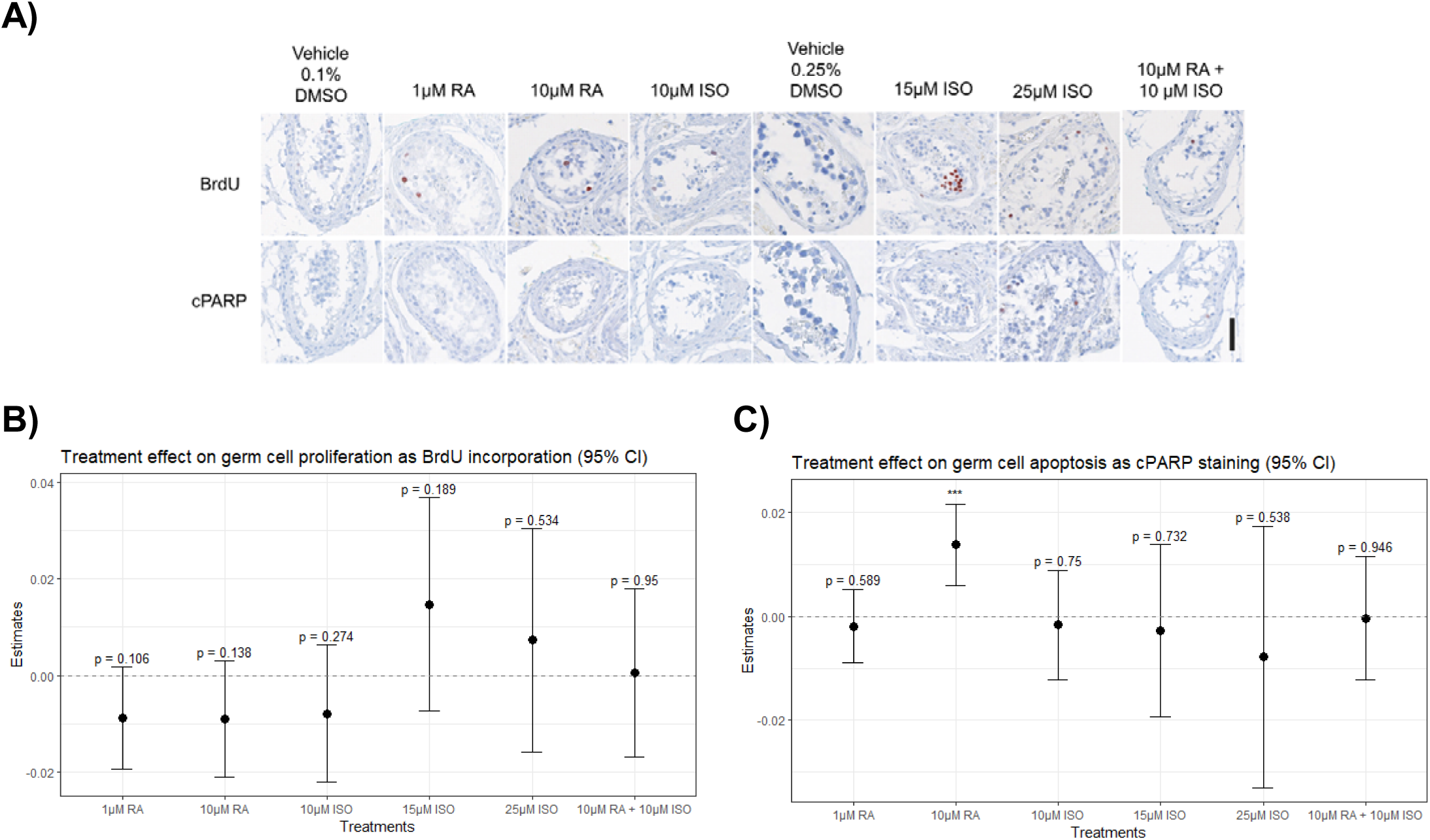
Effect on germ cell proliferation and apoptosis after ISO or RA exposure in cultured human testis tissue. A) Upper panel show IHC staining with incorporated BrdU (Proliferation marker), and lower panels with cPARP (Apoptosis marker). The columns of the panels represent either vehicle control (with 0.1% DMSO or 0.25% DMSO) or the different treatments for the tissue. All sections are counterstained with Mayer haematoxylin. Tissue from 4 donors was investigated, with two experimental replicates per treatment per donor. Magnification 20x, scale bar corresponds to 100µm. B+C) The effect of RA and ISO treatment concentrations (x-axis) on germ cell B) proliferation and C) apoptosis quantified from IHC photos as number of positive cells per tubuli (n=4). The estimates (y-axis) are created with multiple linear regression to their corresponding vehicle control (either 0.1% or 0.25% DMSO). CI of 95%. Dashed line represents the line of null effect. *p<0.05, **p<0.01, ***p<0.001. Abbreviations: BrdU, 5-bromo-2’-deoxyuridin; cPARP, cleaved-poly-polymerase; ISO, Isotretinoin; RA, Retinoic acid.

To investigate whether ISO induced apoptosis, apoptotic cells were identified by c-PARP staining. Few c-PARP-positive cells were detected and quantified in the tissue fragments (Figure 3a, lower panel). The fraction of c-PARP-positive cells relative to all presumed germ cells in each tubule was quantitatively evaluated, and treatment effects are presented relative to the corresponding vehicle control (Figure 3c). No treatment effects were observed on the fraction of c-PARP-positive cells, except after treatment with 10 µM RA, where a small but significant increase in apoptotic cells was found.

### ISO can inhibit the action of RA on RARα activity

To investigate the potential mode of action of ISO, we examined if ISO could inhibit RA-induced RARα activation using the human RARα reporter (Luc)-HEK293 cell line.

As expected, our experiments showed that ISO by itself increased the activity of the RARα receptor in a concentration dependent manner (Figure 4) as in (24). Addition of RA in increasing concentrations to the cells treated with the two highest concentrations of ISO (1440 nM and 195 nM) led to almost no further increase in RARα activity (Figure 4), whereas subsequent addition of RA to cells treated with 12nM ISO (equivalent to EC50 on RARα) led to a concentration dependent increase in response, ultimately reaching an E_max_ value comparable to in cells not pretreated with ISO. However, the EC50 value for RA was shifted to a higher concentration when cells were pretreated with 12 nM ISO compared to in cell without pretreatment with ISO (EC50 = 7.65 µM vs. 2.92 µM, respectively).

**Figure 4:**
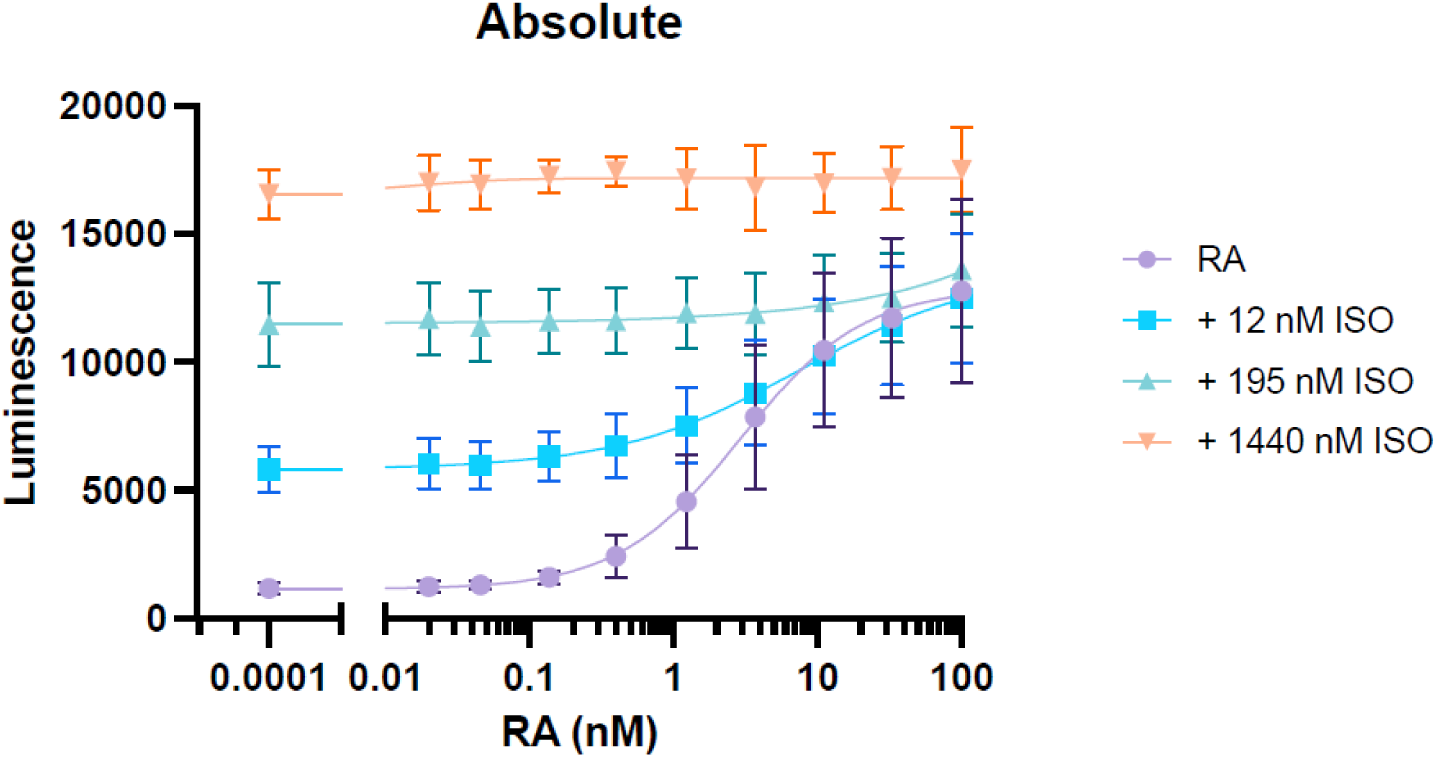
RARα activity after exposure to RA and ISO at different concentrations. Absolute values of luminescence (y-axis) as a measure for retinoic acid alpha (RARα) activity with exposure to first either 12 nM (square), 195 nM (triangle), or 1440 nM (reverse triangle) ISO and RA at increasing concentrations (0.15 pM-100 nM) (x-axis). Graphs represent pooled means from three independent experiments (n = 3).

## Discussion

In this multi-model study integrating human cohort data with *ex vivo* and *in vitro* experimental approaches, we investigated how ISO exposure affect male reproductive function. In a cohort of otherwise healthy young men, ISO-use was associated with higher circulating estradiol concentrations, reduced FSH levels, and a tendency of lower sperm counts. These observations were complemented by altered hormone secretion in cultured human testis tissue exposed to ISO. However, ISO did not affect germ cell dynamics in the *ex vivo* model, suggesting that the observed tendency toward reduced sperm parameters in the cohort cannot be explained by direct short-term effects on germ cells. Importantly, the *ex vivo* model does not fully recapitulate human spermatogenesis, and meiotic processes were not directly assessed, why the results may not reflect testicular function *in vivo*. Finally, ISO was also found to compete with RA for RARα activation *in vitro*, showing that ISO can both activate RARα on its own and inhibit RA-mediated activation of RARα (Figure 4).

Previous human studies of ISO-treated men have generally reported unchanged or increased sperm counts, alongside no or small increases in testosterone and tendencies toward reduced FSH and inhibin B (10,12,15,17,20,21,47,48). In contrast, animal studies report reduced sperm counts and testosterone levels following ISO exposure, with no consistent effects on FSH (7,13,18). In our study, the reduction in FSH aligns with human data but not animal models, whereas the tendency toward lower sperm counts resembles findings from animal studies but contrasts with previous human reports. In the *ex vivo* testis model, ISO exposure increased testosterone secretion and decreased DHT, which resembles findings in some human studies, but not in the animal studies. Data on estradiol in male ISO-users are limited, whereas ISO use in women has been associated with reduced estradiol levels after cessation (48,49). In contrast, we observed slightly elevated estradiol levels in the men after ISO-exposed. Taken together, these discrepancies across studies likely reflect differences in study design, population characteristics, exposure timing and concentrations, as well as potential sex- and tissue-specific endocrine regulation and steroid metabolism. These differences may be further amplified by highly individual regulation of retinoid signaling (50).

Retinoids regulate gene expression via RARs, although the exact mode of action of ISO in *acne vulgaris* remains incompletely understood. While ISO can activate RARα on its own (24), it can also be metabolized to RA (51) and thereby activate RARα. Furthermore, it has been shown that ISO affects changes in expression of genes containing retinoic acid receptor response elements (RAREs) (52), including activation of the transcription factors STRA8, p53, and FoxO1 (53). Our *in vitro* data confirm that ISO can directly activate RARα and show that it can also inhibit RA-induced RARα activation. This means that any spaciotemporal regulation by RA may be altered in the presence of ISO. Given that RA signaling plays a central role in spermatogenesis, where disruption cause infertility, even subtle perturbations may consequently have persistent negative downstream effects on male reproductive function.

The observed hormonal patterns in the ISO-users, i.e., lower FSH combined with a higher inhibin B/FSH ratio and slightly elevated estradiol, suggests altered regulation of the hypothalamic– pituitary–gonadal (HPG) axis. Both inhibin B and estradiol exert negative feedback on FSH secretion, and the increased inhibin B/FSH ratio is consistent with this interpretation. In the *ex vivo* testis model, ISO exposure neither increased estradiol nor inhibin B (but decreased at the highest ISO concentrations or in combination with RA), indicating that testicular effects alone cannot explain the systemic hormonal changes observed in the ISO-users.

Interpreting these systemic versus local testicular effects, it is important to consider the RA and ISO concentrations *in vivo*. Endogenous plasma RA concentrations are in low nanomolar ranges (reported mean values of 6.7 nM), with low or undetectable levels in the seminal plasma (10), while intratesticular RA concentrations in men with normal and abnormal semen analysis have been reported to 0.37 and 0.23 pmol/gram tissue, respectively (54). In ISO treated men, plasma concentrations of ISO have been reported to reach a C_max_ of 1.34-1.44 µM (46,55), with lower concentrations (7-22nM) discovered in the seminal plasma (10). Our data suggest that at reported RA and ISO plasma concentrations there are stark differences in the RA responses observed at the receptor level, in the presence and absence of ISO. Furthermore, it has been shown that even low nanomolar ISO concentrations (3nM) can induce transcriptional responses in testis tissue (56), underscoring that ISO may exert effects well below its C_max_. Together, this indicate that even subtle alterations in retinoid signaling may have biological relevance.

Building on our collected findings, we propose a hypothetical model of ISO action on male reproductive function (Figure 5). In this model, ISO interferes with retinoid signaling by modulating RAR activity, with effects that may vary depending on endogenous RA levels in different tissues. In the testis, ISO exposure increased testosterone levels in the *ex vivo* model, but this may well involve targets other than RAR. Outside the testis, ISO may influence reproductive hormone metabolism, e.g. via increased aromatase activity, as seen for other retinoids (57), which could contribute to the elevated estradiol observed in the ISO-users of cohort. Increased estradiol may in turn contribute to suppress FSH through negative feedback on the HPG-axis.

**Figure 5:**
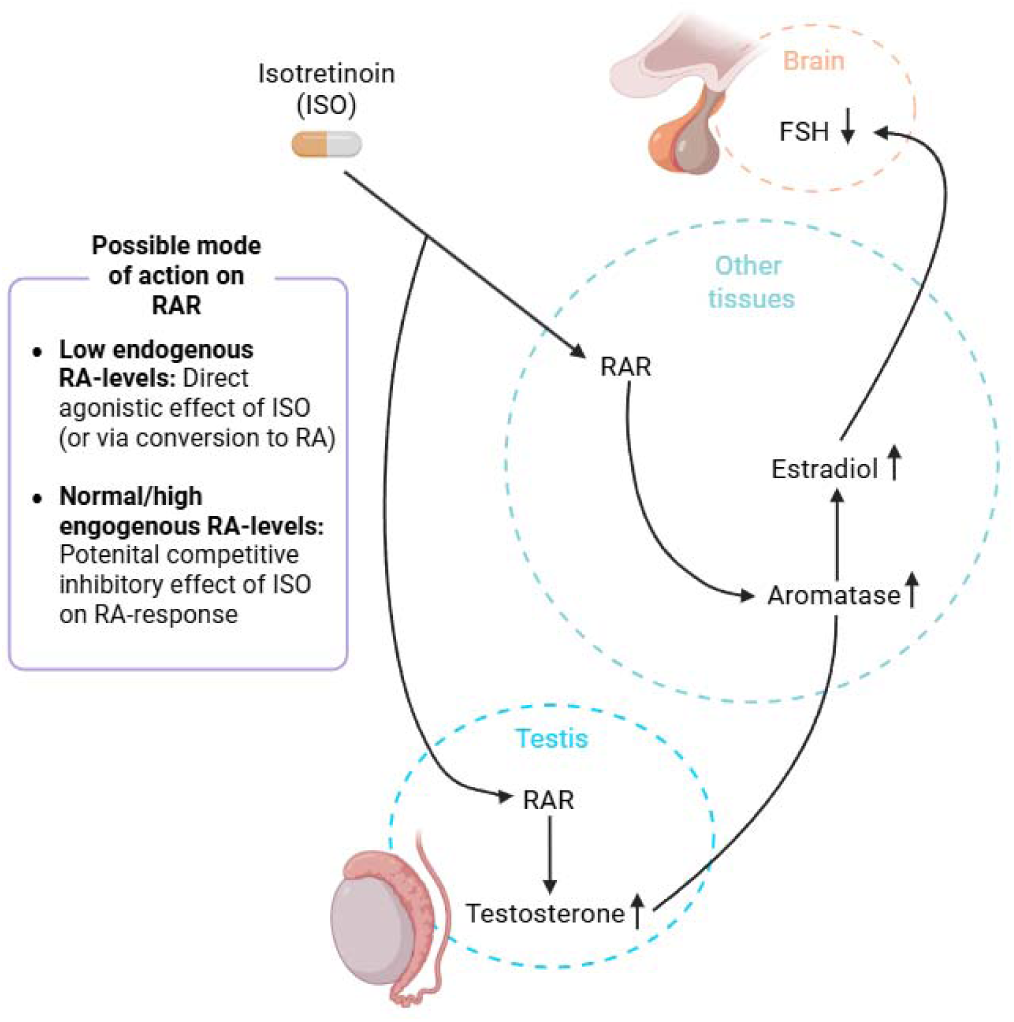
Proposed model for isotretinoin’s effect in male reproductive function. Proposed model of isotretinoin (ISO) action on male reproductive function. ISO may modulate retinoic acid receptor (RAR) activity depending on endogenous RA levels. In the testis, ISO exposure was associated with increased testosterone secretion in *ex vivo* cultures, although the direct mechanism is unclear. In other tissues, ISO may increase steroid metabolism, contributing to increased estradiol levels. Elevated estradiol may suppress FSH through feedback on the HPG-axis. The model is based on integration of cohort, *ex vivo*, and *in vitro* findings and should be interpreted as hypothetical. Figure created using biorender.com.

Although hormone concentrations and semen parameters remained within clinical reference ranges, the consistent direction of the observed changes, even among prior ISO-users, raises the possibility of negative and potentially persistent effects on male reproductive function. Similar long-term effects of retinoids have been reported in male murine studies (13,58), possibly reflecting sustained changes in retinoid-signaling that could be explained by epigenetic mechanisms (59). Other commonly used and otherwise considered safe drugs, such as ibuprofen (60) and selective serotonin reuptake inhibitors (SSRIs) (61), have also been shown to affect male reproductive parameters, although typically only with transient effects.

It is noteworthy, that ISO has been proposed as a potential treatment for male infertility based on short-term improvements in semen parameters in selected patient populations (62). However, these findings are not based on randomized trials and may apply only to specific subgroups. In contrast, our data from a healthy unselected population do not support general beneficial effects of ISO on male reproductive function, suggesting that the positive treatment responses are likely individual and may depend on variability in retinoid signaling and endogenous RA levels.

In conclusion, ISO use was associated with subtle but persistent changes in reproductive hormones in healthy young men, as well as in cultured human testis tissue, and with trends of reduced semen parameters. Our findings raise the possibility that ISO may exert persistent effects on male reproductive function. Further studies are required to confirm these observations in randomized clinical trials.

## Data Availability

All data produced in the present study are available upon reasonable request to the authors. Access to the study cohort data and associated register data is subject to relevant ethical and legal approvals, including permission from the Ethics Committee of the Capital Region of Denmark.

## Author contribution

A.R. and F.B. initiated and designed the study. S.K. contributed to the initial conceptualization of the project. L.P. and N.J. contributed to DYMS cohort data collection, and F.B. and L.P. analyzed these data with interpretation from all three. F.B., H.F., A.JU., and A.JØ. contributed to the *ex vivo* testis culture experiments through data collection, experimental guidance, and data analysis. A.K.R. and T.J. contributed to the *in vitro* reporter gene assay through data collection, experimental guidance, and data analysis. H.C.R. contributed clinical dermatological expertise and interpretation of the findings. L.M. contributed pharma epidemiological expertise and interpretation of the results. F.B. and A.R. drafted the manuscript. All authors contributed to interpretation of the results and the revision of the manuscript for important intellectual content and approved the final version for submission.

