## Supplementary Materials - S1-S4 for "Isotretinoin use is associated with persistent alterations in male reproductive function"

**S1: Representative D2-40 IHC photos of each donor from the *ex vivo* cultured testis tissue**

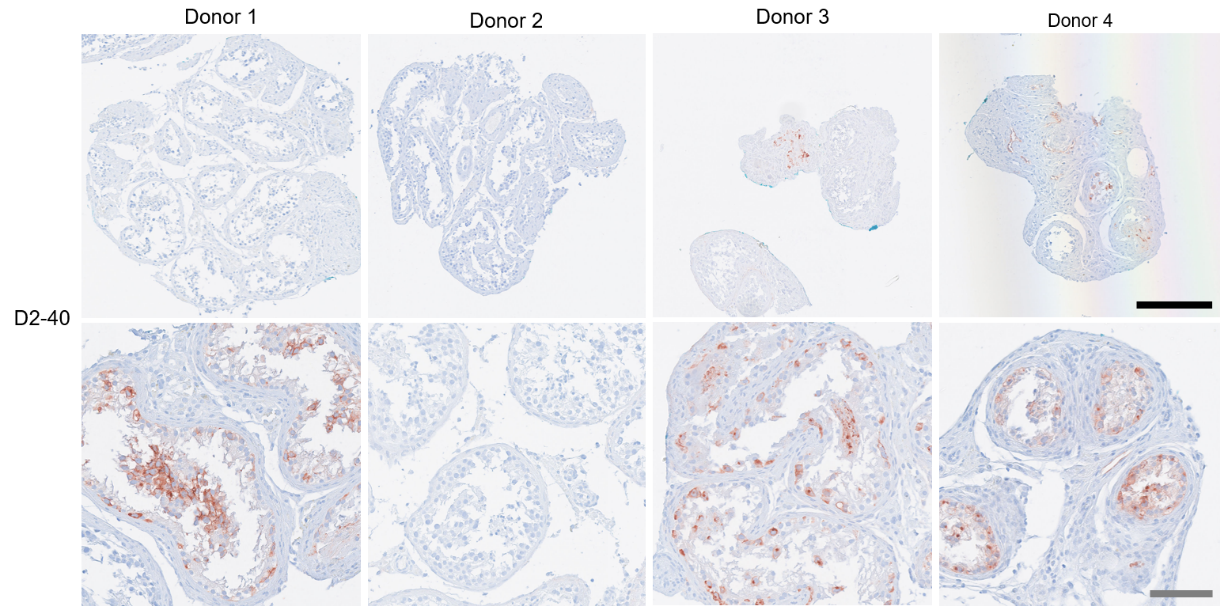

Representative photos of IHC staining with D2-40 (GCNIS marker) showing some tubuli from the donors of testicular cancer patients were GCNIS positive. The columns of the panels are the different donors, the rows show tissue fragments at either magnification of 10x (Upper panel) or 20x (Lower panel). All sections are counterstained with Mayer haematoxylin. Tissue from 4 representative donors was investigated with IHC, with two experimental replicates per treatment per donor. Black/upper scale bar corresponds to 250 µm and grey/lower to 100 µm. A total of 35/88, or 39.77%, of the tissue fragments had 1 or more positive GCNIS tubules, data not shown. Donor 2 (D2) was diagnosed with a spermatocytic tumor and had therefore no positive GCNIS tubles.

**S2: Overview of significant treatment effects on reproductive steroid hormones after ISO and RA exposure in cultured human testis tissue**

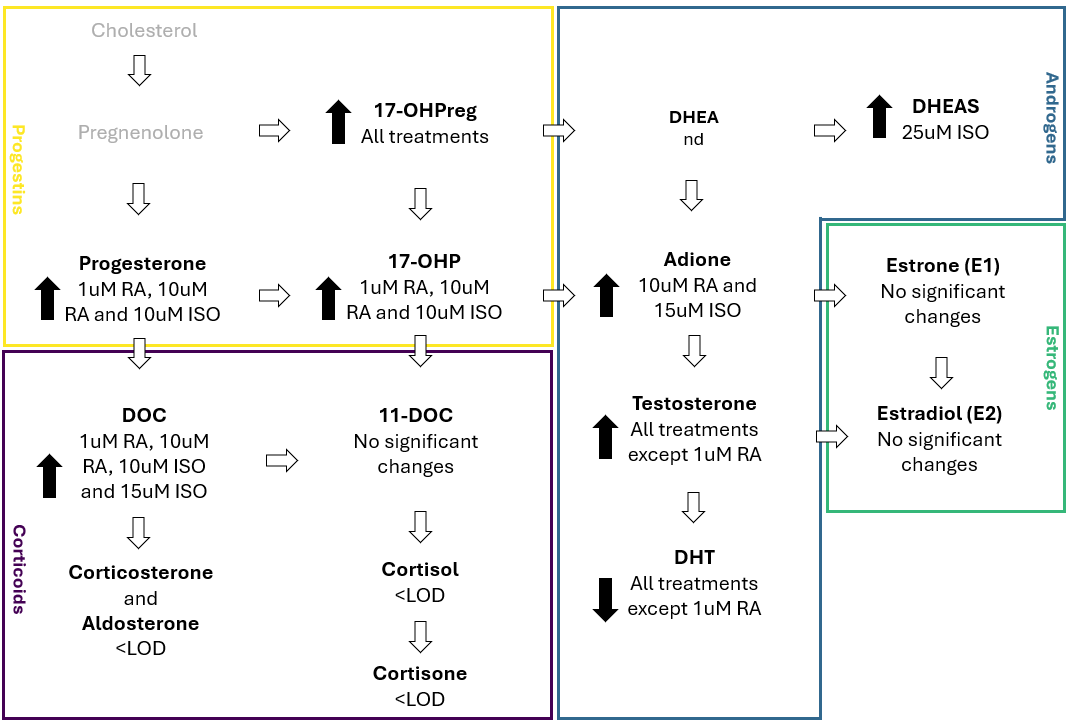

Cholesterol and pregnenolone were not measured. Abbreviations: <LOD, below limit of detection; nd, not detected; DOC, 11β-deoxycosterone; 17OHPreg, 17α-hydroxypregnenolone; 17-OHP, 17α-hydroxyprogesterone; 11DOC, 11β-deoxycortisol; DHEAS, dehydroepiandrosterone sulfate; Adione, androstenedione; DHT, dihydrotestosterone.

| **S3: Effect on the remainder steroid hormones after ISO or RA exposure on cultured human testis tissue** | |
| --- | --- |
| **PROG**  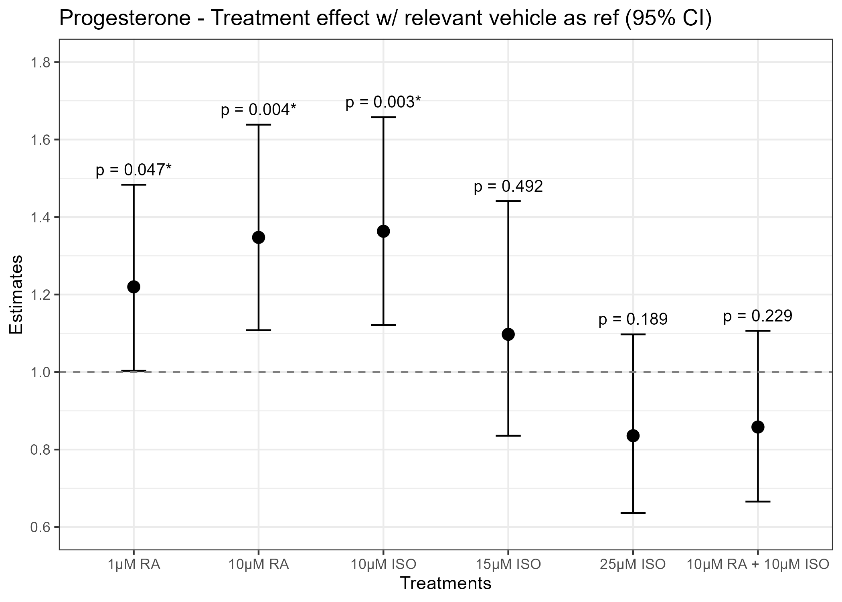 | **DOC** 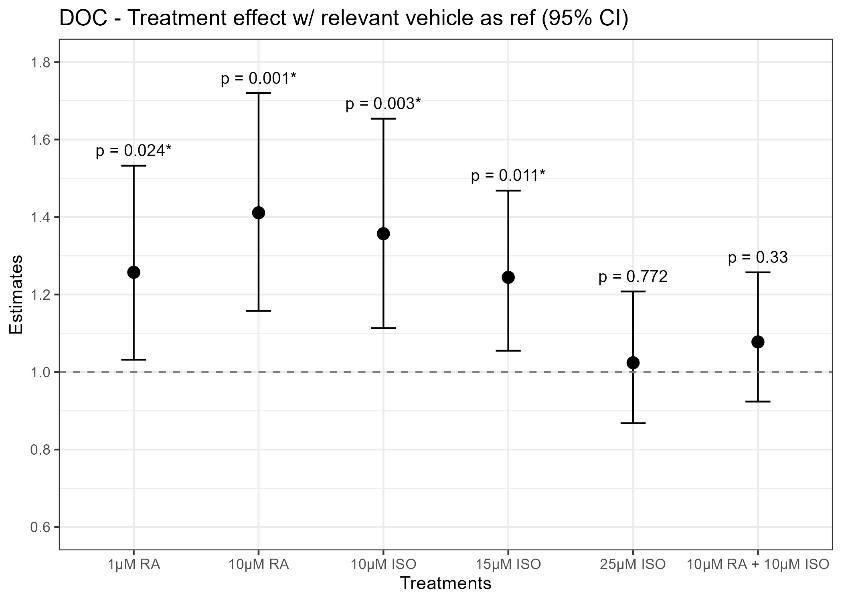 |
| **17-OHPreg**  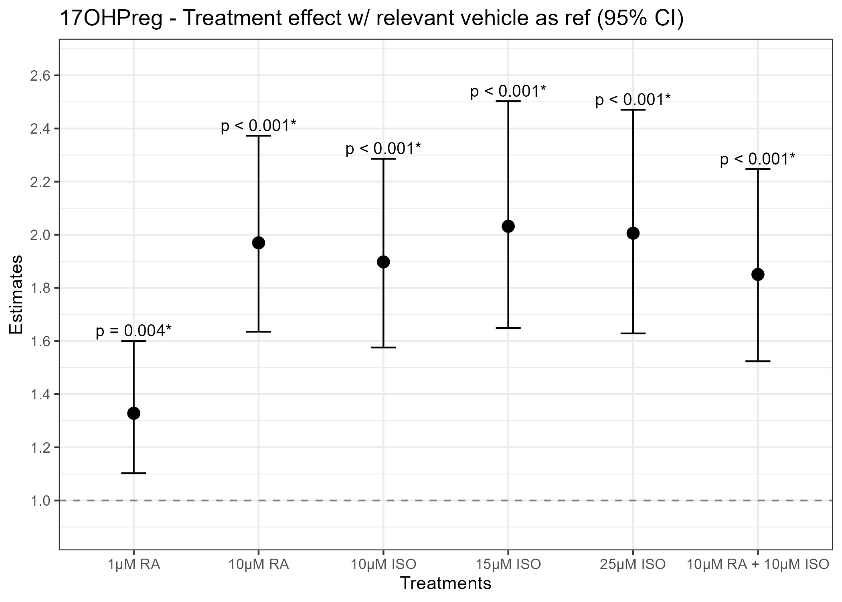 | **17-OHP**  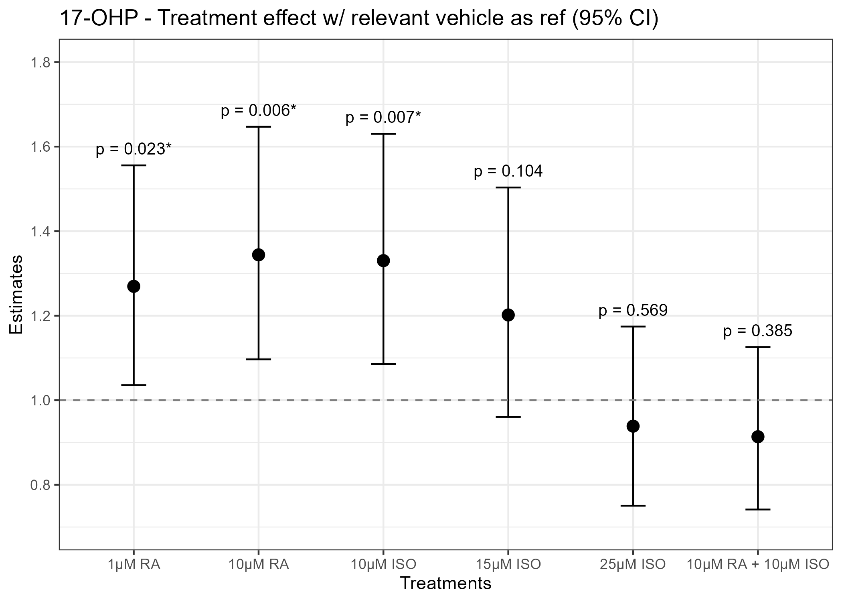 |
| **11-DOC**  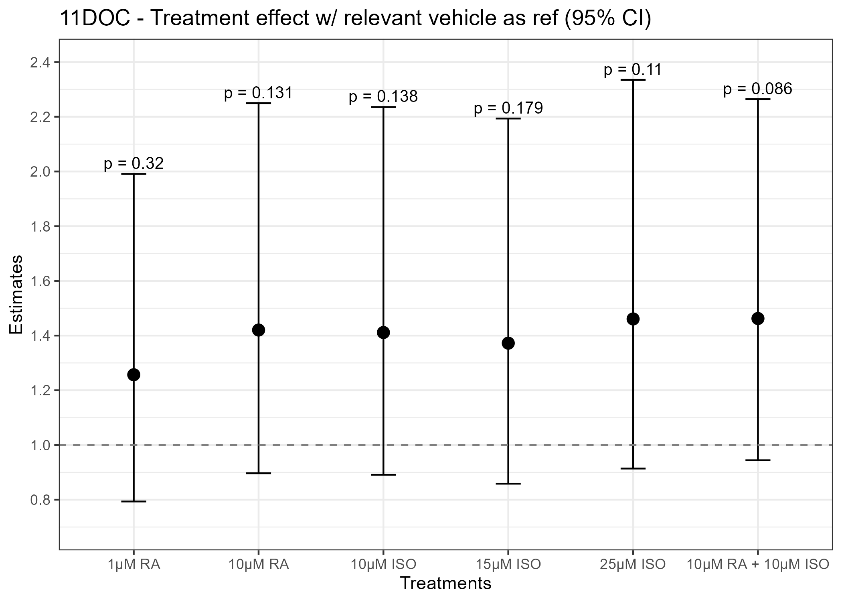 | **Adione**  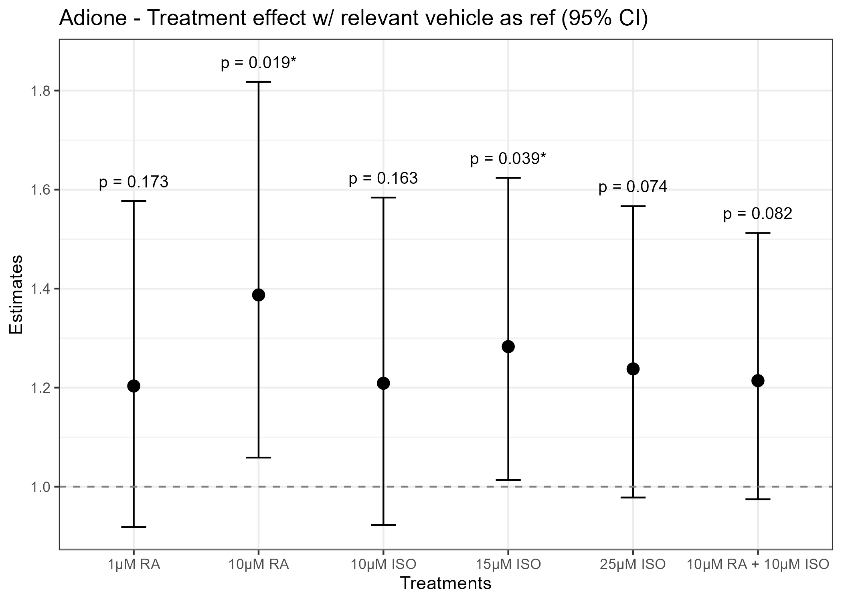 |
| **DHEAS**  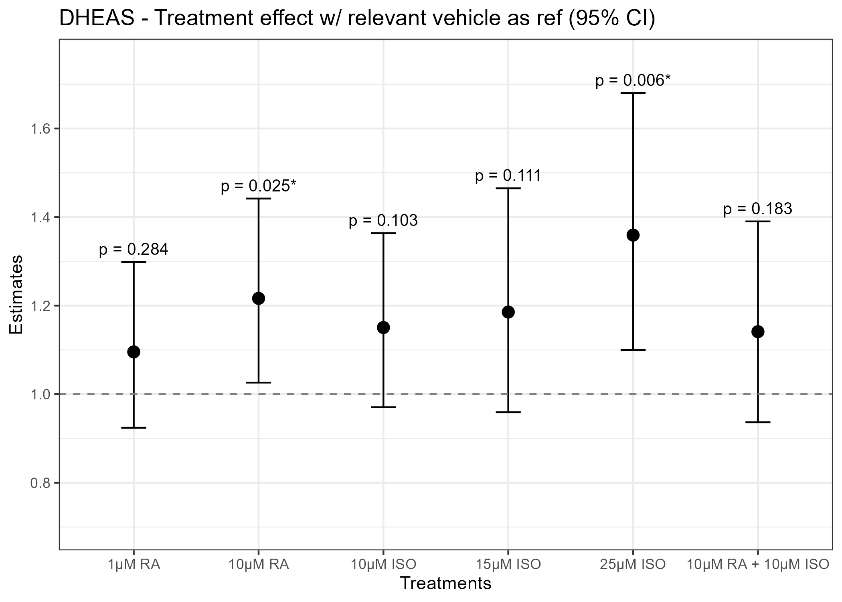 | **Estrone**  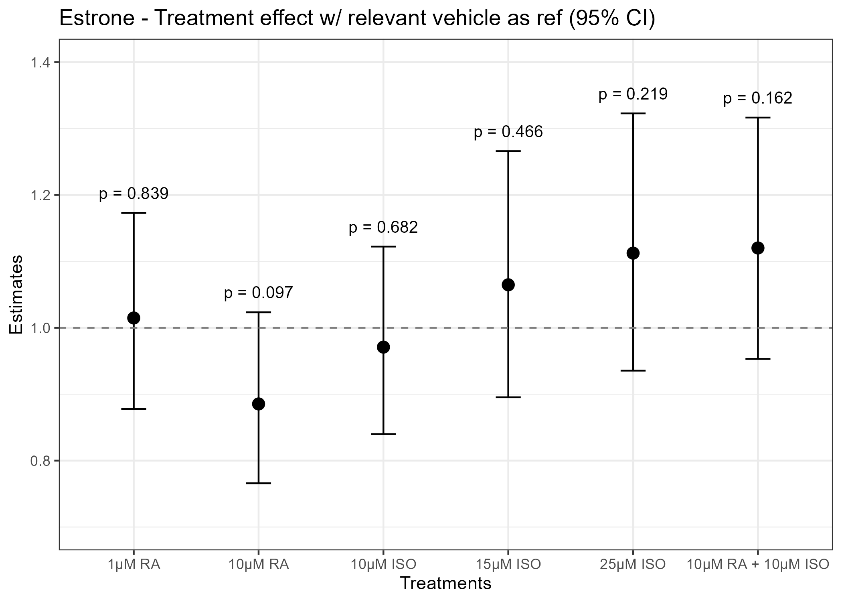 |
| The effect of different RA and ISO concentrations (x-axis) on cultured human testis tissue on the remaining steroid hormones not presented in figure 2 (n=8). The estimates (y-axis) are created with multiple linear regression on log-transformed hormone concentrations to their corresponding vehicle control (either 0.1% or 0.25% DMSO). CI of 95%). Dashed line represents the line of null effect. *p<0.05. Abbreviations: CI, Confidence Intervals; RA, Retinoic Acid; ISO, Isotretinoin; DOC, 11β-deoxycosterone; 17OHPreg, 17α-hydroxypregnenolone; 17-OHP, 17α-hydroxyprogesterone; 11DOC, 11β-deoxycortisol; DHEAS, dehydroepiandrosterone sulfate; Adione, androstenedione; DHT, dihydrotestosterone. | |

**S4: Raw hormone concentrations from cultured human testis tissue after ISO and RA exposure**

**PROG**

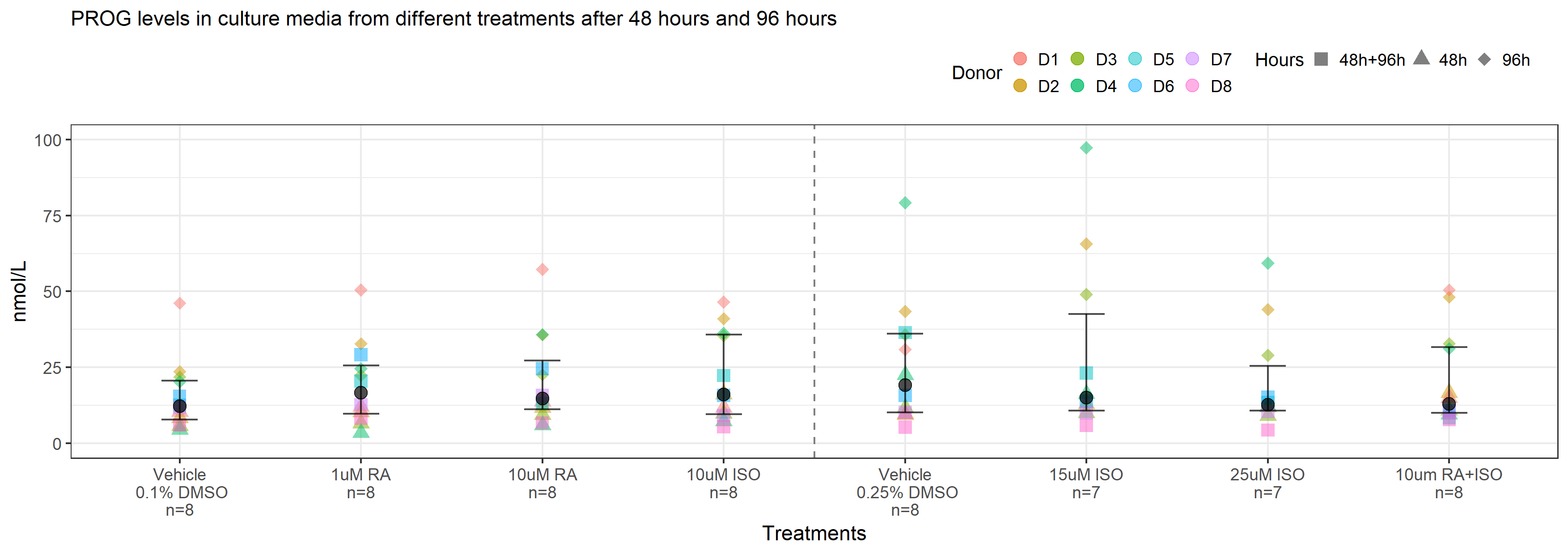

**DOC**

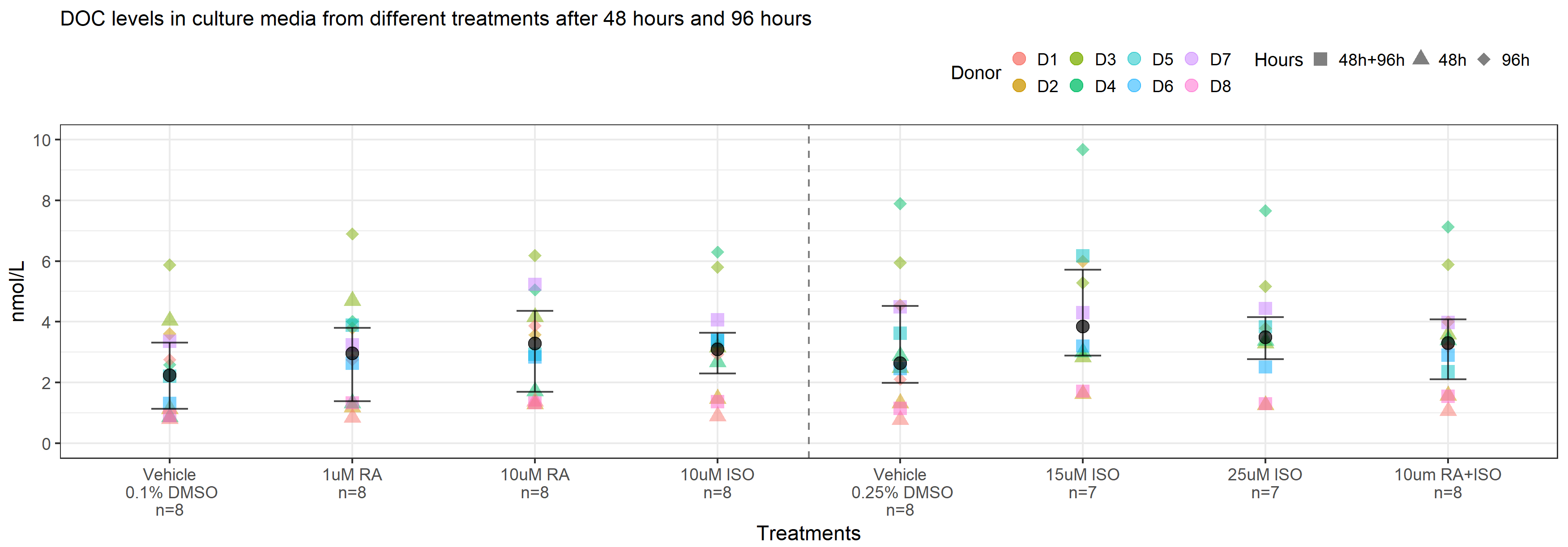

**17-OHPreg**

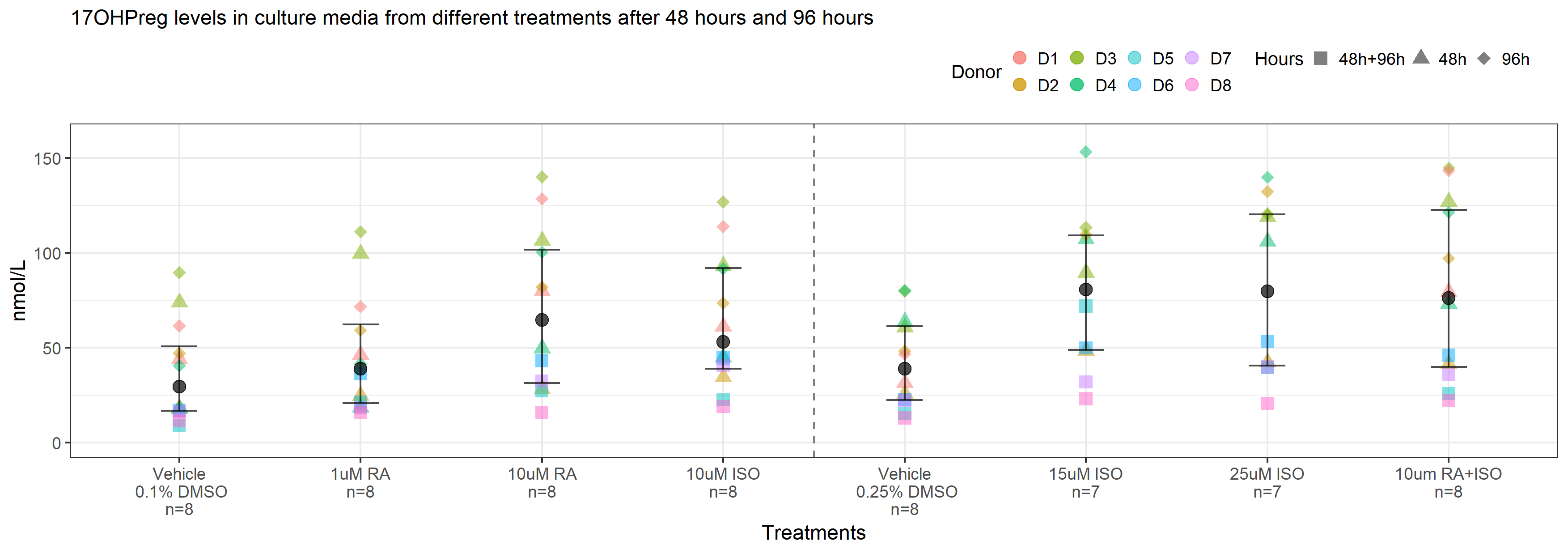

**17-OHP**

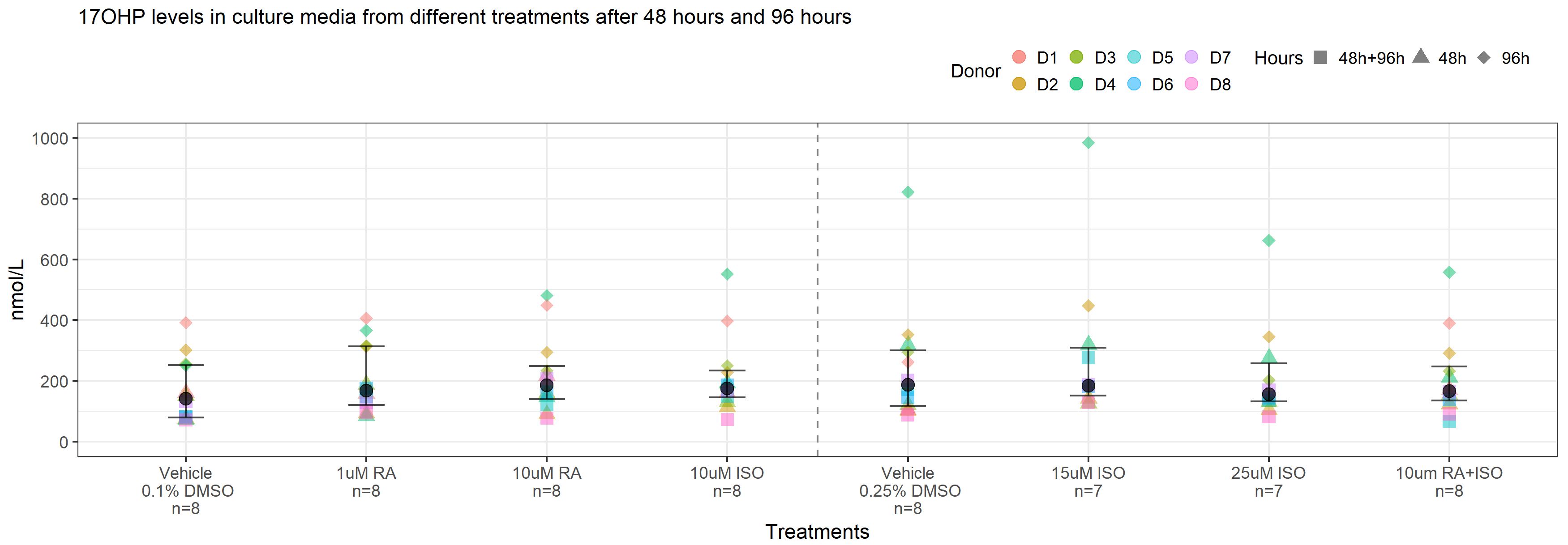

**11-DOC**

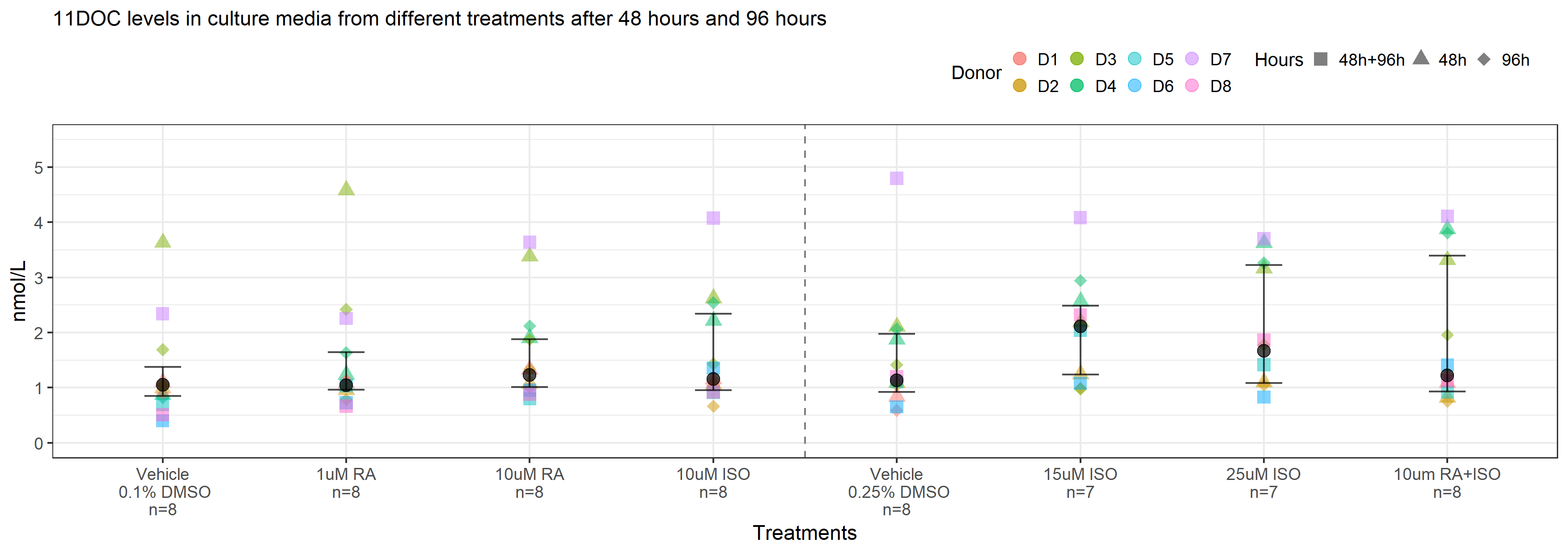

**Adione**

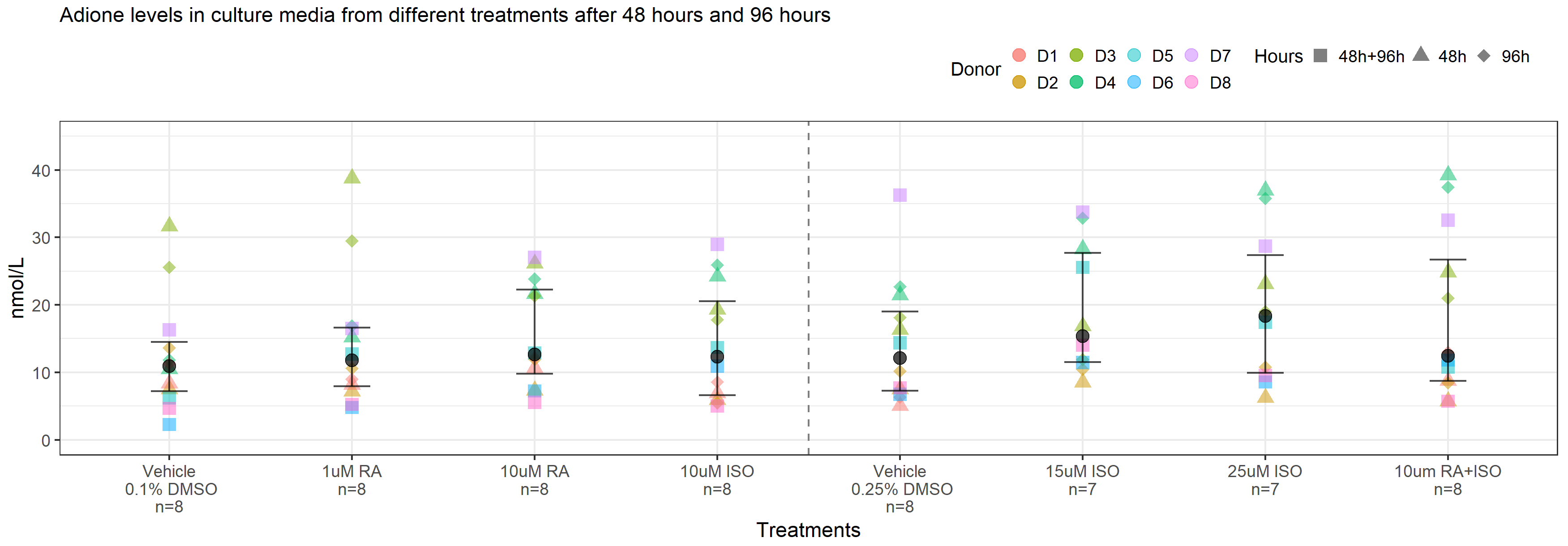

**Testosterone**

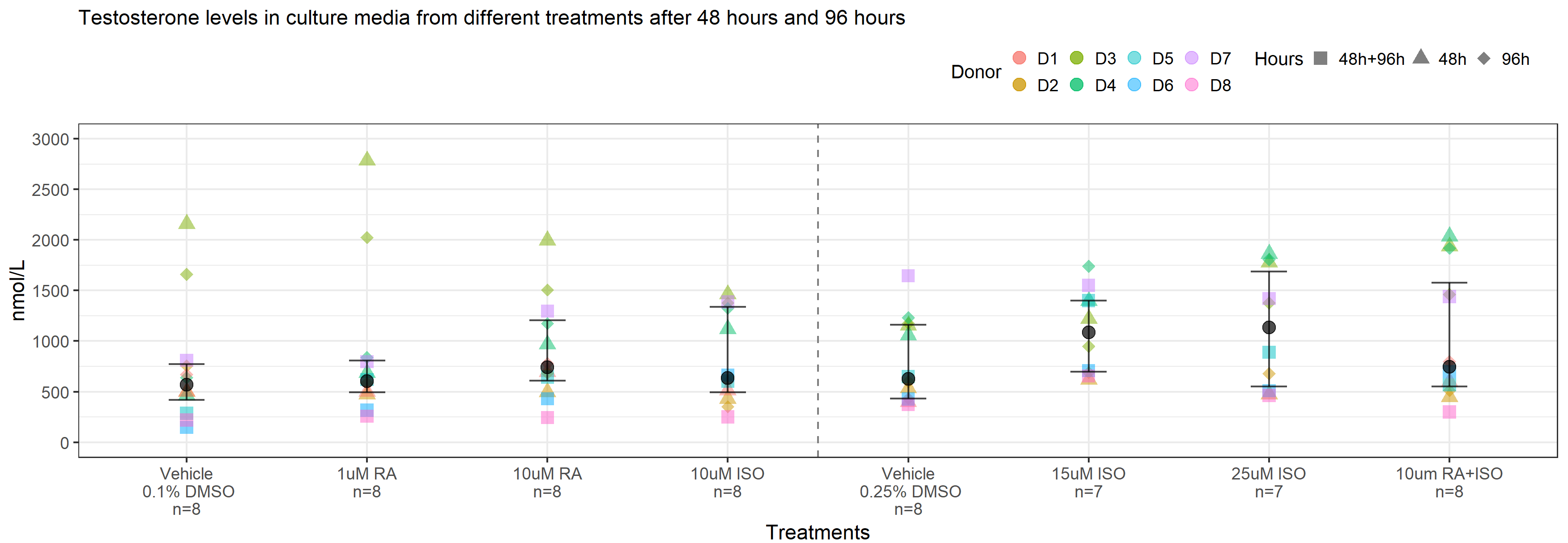

**DHT**

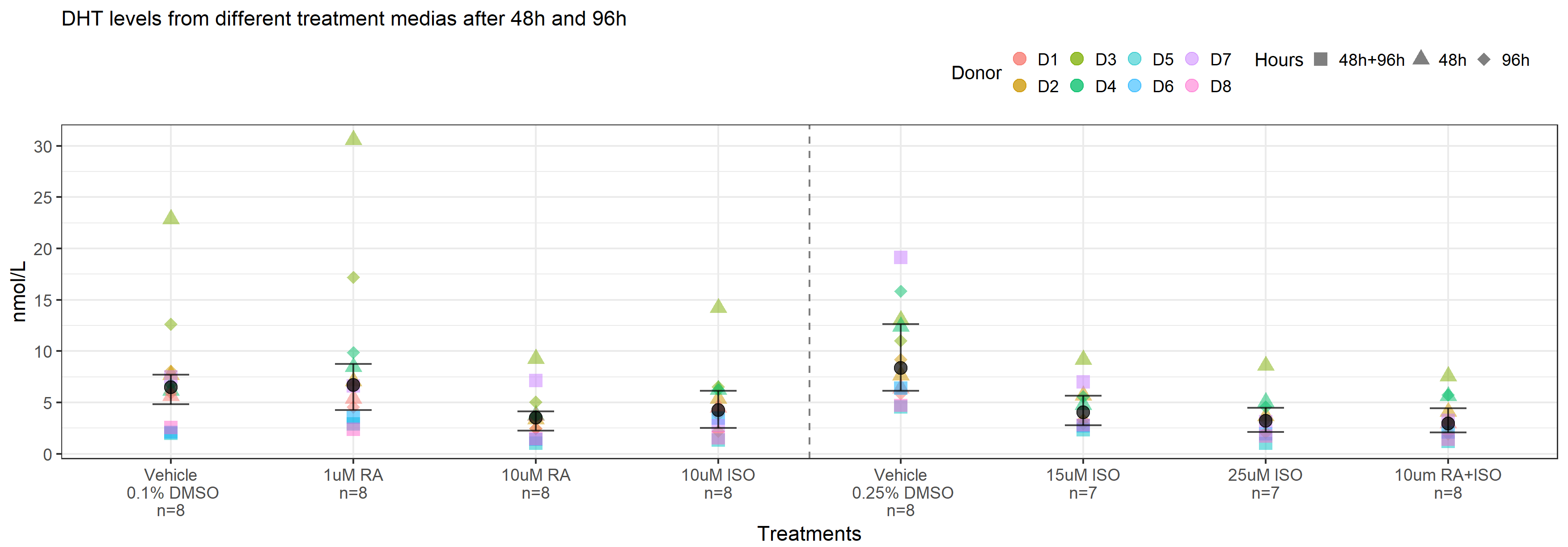

**DHEAS**

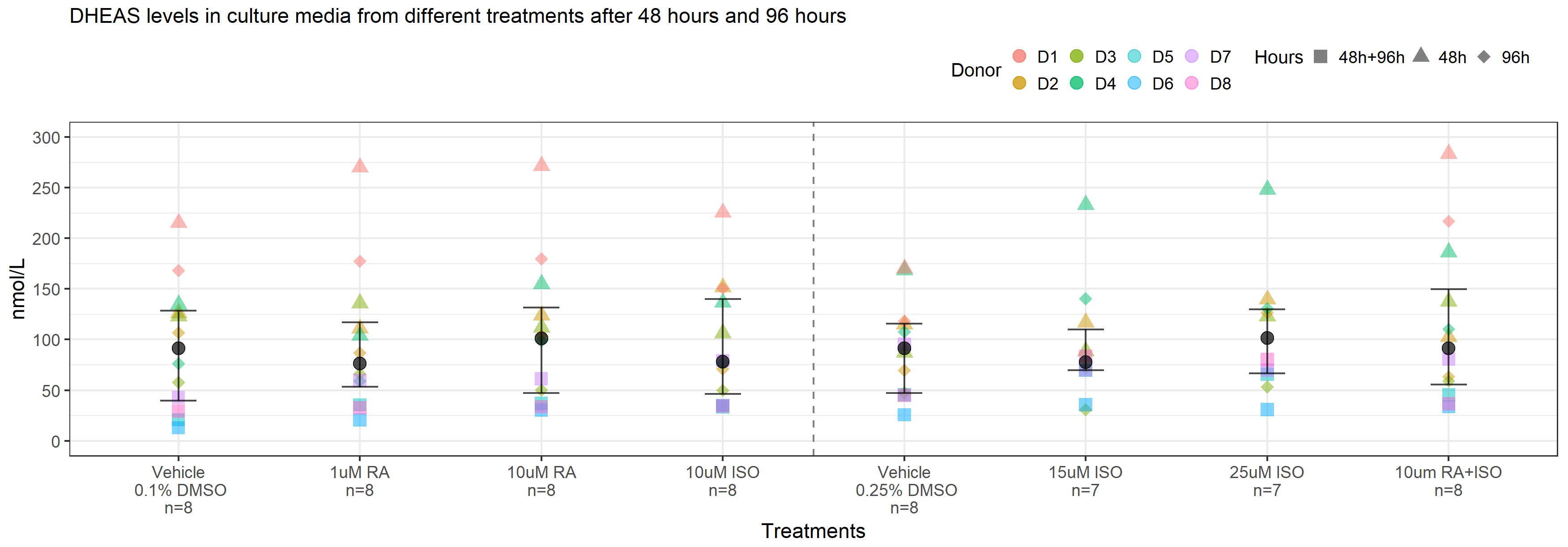

**Estrone (E1)**

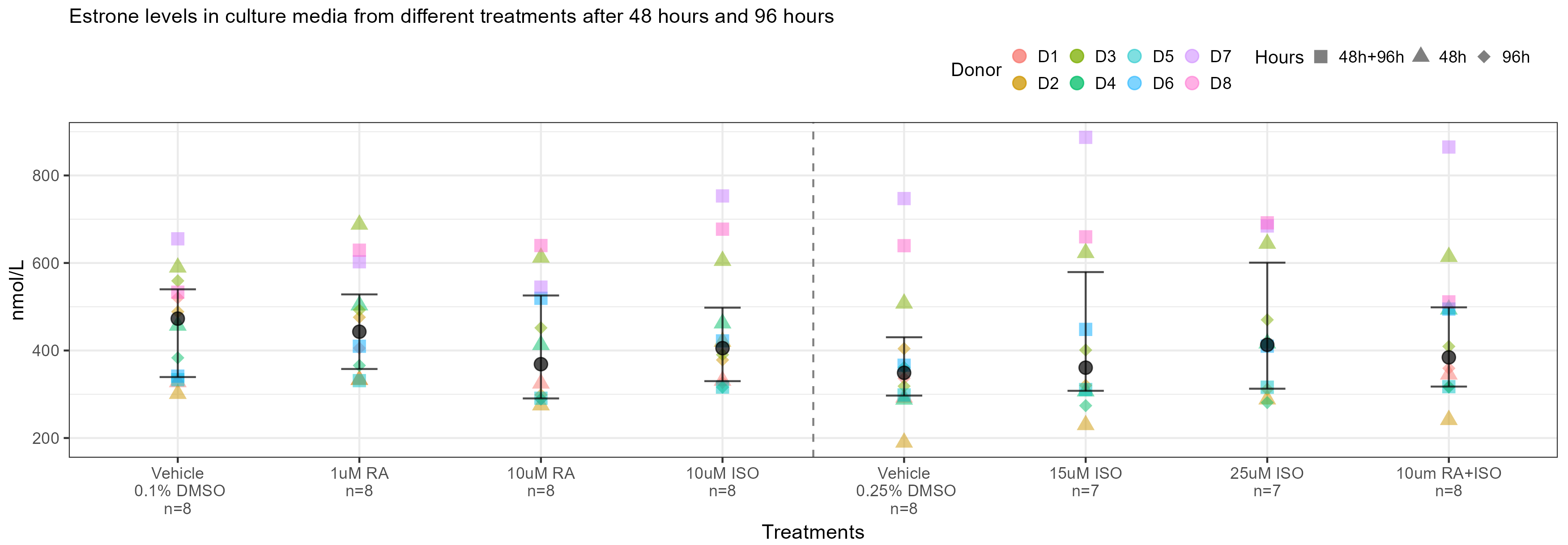

**Estradiol (E2)**

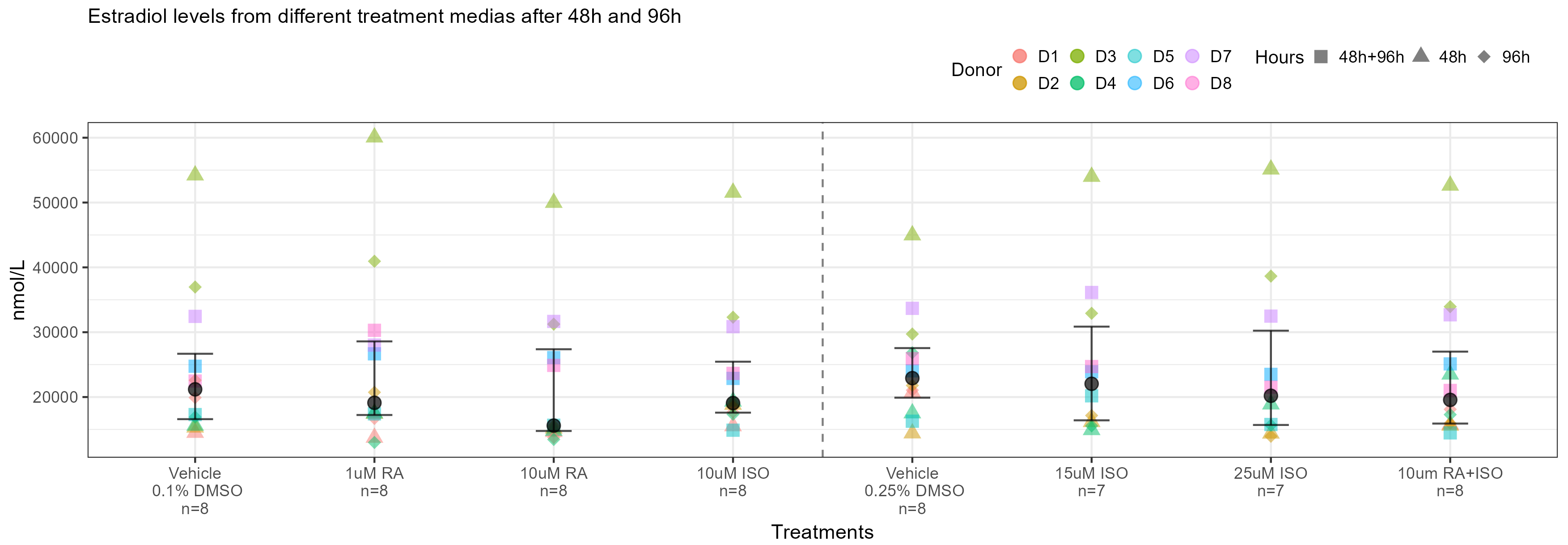

**Inhibin B**

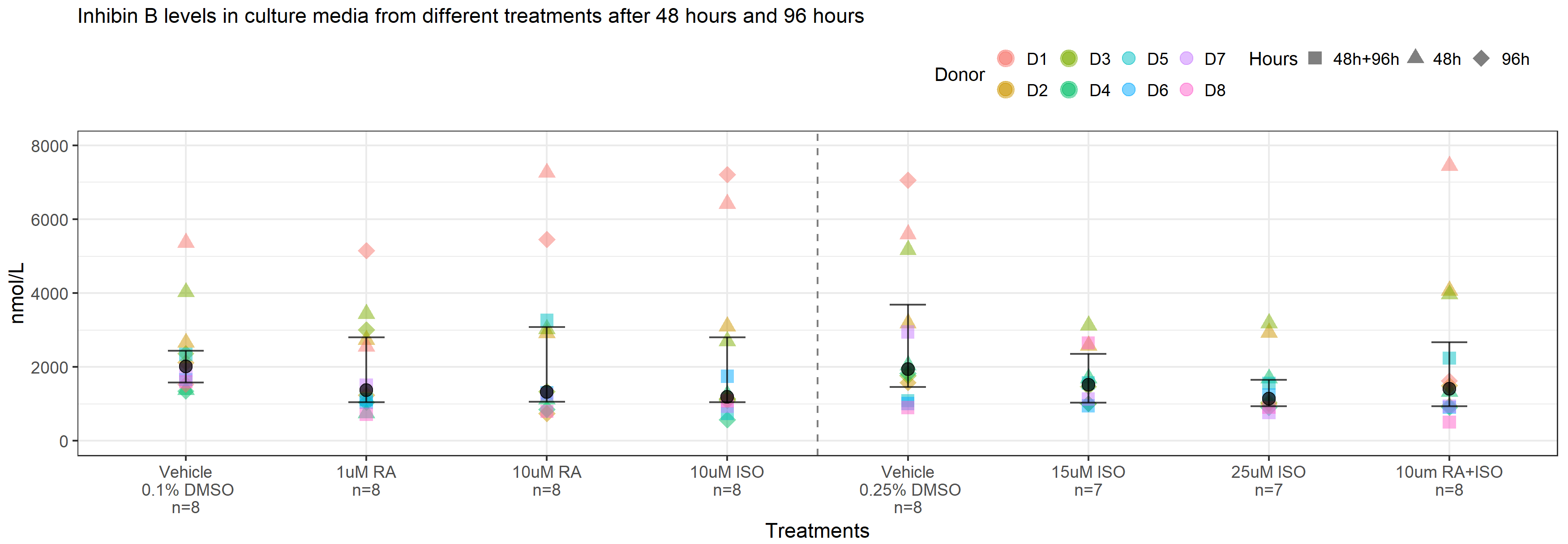

Raw hormone concentrations (y-axis) measured in the treatment media in vehicle (0.1% DMSO or 0.25% DMSO) or after exposure to different RA and ISO concentrations (x-axis) in cultured human testis tissue. The colors represent the different testicular cancer patients (n = 8 or n=7). Circles are measured hormone concentrations after 48 hours and triangles are after 96 hours. The dashed line divides the different treatments in two groups, where treatments on the left have the vehicle of 0.1% DMSO as control/reference and treatments on the right have the vehicle of 0.25% DMSO as control/reference. Abbreviations: RA, Retinoic Acid; ISO, Isotretinoin; DOC, 11β-deoxycosterone; 17OHPreg, 17α-hydroxypregnenolone; 17-OHP, 17α-hydroxyprogesterone; 11DOC, 11β-deoxycortisol; DHEAS, dehydroepiandrosterone sulfate; Adione, androstenedione; DHT, dihydrotestosterone.
